# A Solution to the Kermack and McKendrick Integro-Differential Equations

**DOI:** 10.1101/2022.04.28.22274442

**Authors:** Ted Duclos, Tom Reichert

## Abstract

In this manuscript, we derive a closed form solution to the full Kermack and McKendrick integro-differential equations (Kermack and McKendrick 1927) which we call the KMES. We demonstrate the veracity of the KMES using independent data from the Covid 19 pandemic and derive many previously unknown and useful analytical expressions for characterizing and managing an epidemic. These include expressions for the viral load, the final size, the effective reproduction number, and the time to the peak in infections. The KMES can also be cast in the form of a step function response to the input of new infections; and that response is the time series of total infections.

Since the publication of Kermack and McKendrick’s seminal paper (1927), thousands of authors have utilized the Susceptible, Infected, and Recovered (SIR) approximations; expressions putatively derived from the integro-differential equations to model epidemic dynamics. Implicit in the use of the SIR approximation are the beliefs that there is no closed form solution to the more complex integro-differential equations, that the approximation adequately reproduces the dynamics of the integro-differential equations, and that herd immunity always exists. However, the KMES demonstrates that the SIR models are not adequate representations of the integro-differential equations, and herd immunity is not guaranteed. We suggest that the KMES obsoletes the need for the SIR approximations; and provides a new level of understanding of epidemic dynamics.

## Introduction

Modern epidemiological modeling has its roots in the Kermack and McKendrick epidemic model first published in 1927 (Kermack and McKendrick 1927). Since its publication, well over 10,000 authors have referenced this paper and used it as a foundational starting point. Throughout this vast literature, three basic tenets are held to be true: 1) There are no known or published closed-form solutions to the full set of Kermack and McKendrick’s integro-differential equations; 2) An approximation, known as the SIR (Susceptible, Infected, Recovered) model and its variants, are accepted as reasonable representatives of the full equations; and 3) the final number of uninfected individuals is always greater than zero (i.e., it is not possible for everyone to become infected). This last property is referred to as “herd immunity”.

Despite the durability of these associations, in this manuscript, we explicate that a closed-form solution to the equations can be derived; and based on this solution, we demonstrate that the arguments and mathematics used to justify the use of the SIR model, as well as those advanced to prove that herd immunity exists, rest upon faulty assumptions. As a preview to our approach, in this introduction, we present an outline for this solution and use this to highlight some of the inadequacies in the reasoning that supports the existence of herd immunity and other, improbable, properties of the SIR model.

### General Outline of the Solution

We begin by defining several quantities which describe the epidemic dynamics. These are: *N*_*p*_, the total population of people who can become infected during the epidemic; *S*(*t*), the subpopulation that has not yet become infected; *N*(*t*), the subpopulation that is currently or has previously been infected; *I*(*t*), the infectiousness of individuals within *N*(*t*); and R(*t*), the total recovery of individuals within *N*(*t*). The relationships between these quantities are further clarified by noting that *S*(0) is the number of uninfected people in *N*_!_ at the epidemic start, *S*(∞) is the number of uninfected people at the end of the epidemic, *I*(0) is the number of infectious individuals at the epidemic start; and therefore, *N*_!_ = *S*(*t*) + *N*(*t*), *N*(*t*) = *I*(*t*) + *R*(*t*), and *N*(0) = *I*(0). We further assume that for our modelling purposes, *N*_!_ is a constant and that once infected, people cannot become reinfected.

We then write the following two relationships,

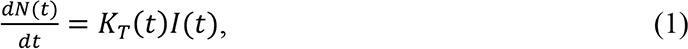

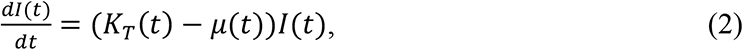

where *K*_*T*_(t) is a function describing the rate of new infections and *μ*(*t*) is the recovery rate of the infected people. Equation 1 arises directly from the notion that the currently infectious subpopulation, *I*(*t*), is the sole cause of new infections. In a “systems” view, infections are expressly recognized as the driving force of the epidemic.

Equation 2 is easily solved to find an expression for I(*t*),

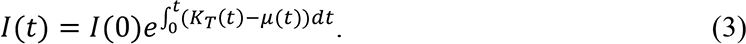

which can be rewritten as,

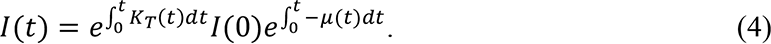

The righthand side of Equation 4 is the decaying step input of the initial infectious individuals, 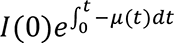, multiplied by the response function, 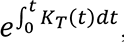, to this input. Corroborating this simple systems view, we see that if there were no recovery (i.e., if *μ*(*t*) = 0 for all time), then as long as *K*_*T*_(*t*) > 0, the number of infectious individuals would grow exponentially.

### Herd Immunity

With Equation 4 in hand, we can now explore several accepted tenets of epidemiological modeling. One generally accepted tenet, the concept of herd immunity, requires the determination of the epidemic’s final size. To see if our model predicts herd immunity, we first use Equations 1 and 3 to determine the time history of the total infections, N(*t*); and find,

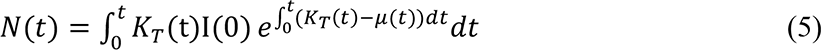

We then note that iff the condition *N*(*t*) = S(0) can occur, *S*(∞) = 0 is possible; and herd immunity is not guaranteed.

By inspection of Equation 5, it is clear there are finite values of *K*_*T*_(t) and *μ*(*t*), which allow N(t) to assume any finite value, including *N*(*t*) = S(0). Therefore, unless there are heretofore unknown or unstated physical restrictions on the range of values for *K*_*T*_(t) and *μ*(*t*), it is plausible that *S*(∞) can be zero; and herd immunity is not guaranteed. This finding is provocative and suggests that we carefully examine the logic used to reach the conclusion that herd immunity, a sine qua non of conventional epidemiological modeling, does always exist.

The conventional approach to deriving an expression for the final size of an epidemic begins with the following summarized Kermack and McKendrick equations (Kröger M and Schlickeiser 2020),

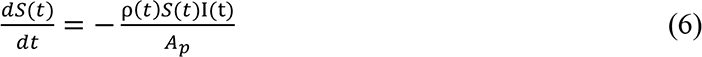

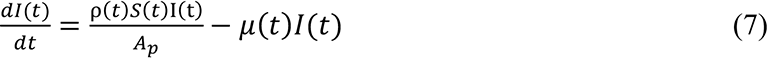

where ρ(*t*) and µ(*t*) are the time varying versions of the parameters that Kermack and McKendrick define respectively as the “rate of infectivity” and “the rate of recovery”, and *A*_*p*_ is the area that encompasses the population.

The conventional final size derivation proceeds by dividing both sides of Equation 6 by *S*(*t*). But, within this early step hides a fundamental assumption. To divide Equation 6 by S(*t*), we must first assume that both S(*t*) and *S*(∞) are ≠ 0 for all time. Thus, the conventional demonstration that *S*(∞) ≠ 0, the essence of herd immunity, begins with the assumption that the conclusion is true.

Adherents of the analysis justify the assumption that S(*t*) and *S*(∞) are ≠ 0 by either explicitly or implicitly assuming and accepting that ρ(*t*) or its equivalent, is finite. Using the further assumption that ρ(*t*) and µ(*t*) are constants, and employing Equations 6 and 7, they then arrive at the following implicit expression for determining the final size (Brauer, et al 2008),

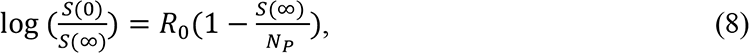

where 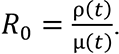. When presented in these analyses, Equation 8 is also accompanied by the statement ρ(*t*) < ∞; and therefore *S*(∞) > 0. There are variations on this theme, but in all cases the conclusion that *S*(∞) > 0 rests on the assumption that ρ(*t*) < ∞.

The critical assumption, that ρ(*t*) < ∞, is supported in these analyses by making one or the other of two physical assertions. One assertion is that ρ(*t*) is the contact rate between the susceptible and infected populations (Brauer 2005) and therefore can never be infinite. In the second assertion, the quantity I(t)ρ(*t*) is referred to as the “force of infection” (Breda, et al, 2021, Diekmann, et al, 2021) which, by its very nature, is assumed to be finite.

However, a simple dimensional analysis of Equation 5 lays bare the historical controversy and contravenes the assertions that *ρ*(*t*) must remain finite. In Equation 6, the units of ρ(*t*) and I(t)ρ(*t*) must be: 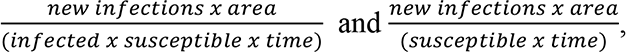 respectively. Therefore, based on their units and their definition, as S(*t*) approaches zero, because each new infection occurs in proportion to an ever-decreasing value of S(*t*), both ρ(*t*) and I(t)ρ(*t*) should be expected to become increasingly large and approach infinity. Beyond these dimensional problems, questions such as upper limits and functionality are simply unaddressed and unanswered in these analyses. The arguments that ρ(*t*) and I(t)ρ(*t*) must remain finite are, therefore, unconvincing.

### Flatten the Curve

Despite the foregoing analysis, in typical analyses that use the SIR model, as a simplifying assumption, ρ(*t*), the rate of infectivity, is replaced in Equations 6 and 7 by the constant *β*. Importantly, the origins of both parameters ρ(*t*) and *β* have their roots in Kermack and McKendrick’s initial assumption that the population is well mixed. The term *S*(*t*)I(t) in Equation 6 quantifies the well mixed population; but since it is absurd to think that everyone in a population could possibly contact everyone else, Kermack and McKendrick introduced ρ(*t*) in an attempt to capture the transmissions occurring due to the actual level of interactions by stating that “…the chance of infection is proportional to the number of infected …, and to the number not yet infected…”; and ρ(*t*) is the time-based parameter of proportionality. The use of ρ(*t*) was intended to capture the dynamics of the disease transmission and the population interactions and this purpose is extended to the use of the constant *β*. In the literature describing the SIR model, *β* is thought of as proportional to and a proxy for the actual interactions.

However, simply multiplying *S*(*t*)I(t) by a parameter, here the constant *β*, is not the proper method to account for the actual number of interactions, because regardless of the value of ρ(*t*) or *β*, the use of the term *S*(*t*)I(t) in Equations 6 and 7 signifies that everyone is still in contact with everyone else. Lower values of *β*, then, merely represent less efficient transmission during those interactions. *β* is not a proxy for the level of population interactions; rather, it is a parameter describing the efficiency of transmission.

We highlight the commonplace misrepresentation of *β* as a proxy for the population interactions because it has led authors to attribute the SIR model phenomenon known as “flatten the curve” to reduced population interactions. Many modelers have shown that lower values of *β* in the SIR model project the peak in new infections to occur later (See, for example, Di Lauro, et al, 2021). Since they believe *β* to be a proxy for the population behavior, they then propose that lower numbers of population interactions will lead to a later epidemic peak. During the Covid-19 pandemic, the belief in this relationship led to many public and political discussions about whether it was more beneficial to shutdown economies and delay the end of the epidemic or to allow the epidemic to peak quickly and end sooner.

Socio-tragically, the “flatten the curve” discussion is based on the incorrect interpretation of the role that *β* plays in the SIR model. The proper interpretation of a lower *β* is that this represents less efficient disease transmission. That the peak is pushed out in time with less efficient transmission is not surprising, but this should not be confounded with lowering the number of population interactions. Contrary to popular belief, as we show in this manuscript, when population interactions are properly considered, lower numbers of interactions significantly shorten the time of the epidemic.

### Coincidence of the Peaks of Infections and New Infections

While the posited phenomena of herd immunity and “flatten the curve” are well known characteristics of the SIR models, a lesser known, but equally important phenomenon is that the SIR model with constant coefficients predicts that the peak in the new cases, 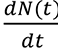, will occur earlier than the peak in infections, I(t). This can be seen by substituting the constants *β* and *γ* for ρ(*t*) and µ(*t*), respectively, in Equations 6 and 7. Then, by first differentiating Equation 6 and setting the quantity 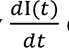 equal to zero, it is clear that 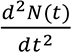 is negative when 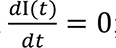; indicating that the SIR model projects that the daily number of new infections will have already peaked before the only source of new cases, the infections themselves, has peaked.

We created Equation 1 from the simple premise that all new infections are caused by, and directly flow from existing infections; that is, apart from an original inoculum, no new infections are introduced from outside the population under consideration. This assertion applies to all models; and, therefore, it should be expected that the projected course of new infections in every model will parallel the course of the infections and peak when the infections peak. Clearly, if *K*_*T*_(*t*) is a constant in Equation 1, in an epidemic described by that equation, the peak in new cases coincides with the peak in infections. In contrast, this is never the case in SIR models.

We have argued above, that in the SIR model with constant coefficients, *β* should only characterize the efficiency of disease transmission. Therefore, one might suspect that the reason new cases peak before the peak in infections in the SIR model is that a hidden phenomenon exists within the SIR model. As we lay out in this manuscript, this hidden phenomenon is equivalent to an assumption of quite implausible population behaviors which have been obscured by the SIR model structure.

### Our Approach

In this manuscript, we use the full Kermack and McKendrick integro-differential equations and an appropriate definition of the population interactions to develop a complete solution, which we refer to as the KMES (**<u>K</u>**ermack and **<u>M</u>**cKendrick **<u>E</u>**quation **<u>S</u>**olution). We validate the KMES by accurately projecting data obtained from the Covid-19 pandemic. Building on this, we derive many useful, new analytical formulas for characterizing and managing an epidemic. Included in these are expressions for the viral load and for both N(*t*) and N(∞).

Lastly, the availability of a closed form solution enables us to closely examine the assumptions behind the SIR epidemiological compartmental models. In that examination, we find implicit, implausible assumptions, which have not been previously appreciated. This last exercise also demonstrates that the conventional imagery of people travelling irreversibly from one compartment to the next, even under the assumption of perfect immunity in recovery, requires significant adaptation.

## Section 1: Solution to the Kermack and McKendrick Equations

(Note: We use the following equation notation, (X, SY-Z), where X is the equation number in the body; and if the equation is used in a supplement, Y is the supplement number and Z is the number of the equation in the supplement). A list of all equations is provided in Supplement 7.

### Definition of terms

Similar to SIR models, the KMES only requires the specification of two parameters: one describing disease dynamics and a second describing the dynamics of population interactions. In the KMES, the disease dynamics are described by *K*_*T*_(*t*), the rate at which the infectious people cause new infections; while the population dynamics are described by *P*_*c*_(*t*), the number of infectable contacts between people within the entire population. We define *P*_*c*_(*t*) with the expression:

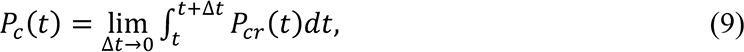

where *P*_*cr*_(*t*) is the rate of infectious contact for the entire population. *P*_*c*_(*t*) is understood to be the instantaneous average of the contacts within the population. Infectious contacts can be fractions of a whole between two contacts; in other words, there is an amplitude associated with every contact and these weighted contacts average to *P*_*c*_(*t*) for the population.

To begin the derivation of the KMES, we first define *I*(*t*) and *R*(*t*) more precisely. In the conventional literature *I*(*t*) is conceptualized as the sub-population that is currently able to advance the epidemic by infecting previously uninfected people; and *R*(*t*) is thought to represent people who have been infected, but because they have recovered, can no longer infect anyone. As the epidemic progresses, the members of the population *I*(*t*) move in a one way sense into the population, *R*(*t*).

To develop the KMES, we defined *I*(*t*) as the infectiousness of the population *N*(*t*) where, by “infectiousness”, we mean the ability to infect others. We define *R*(*t*) as the portion of *N*(*t*) that is unable to infect others and is therefore “not infectious”. In the following three thought experiments, we illustrate how our definitions of *I*(*t*) and *R*(*t*) expand on conventional notions.

In the first thought experiment, we imagine a person with an active infection who has no contacts at all or only with persons who cannot become infected. This person is certainly a member of the population *N*(*t*); but since they cannot infect anyone, they cannot advance the epidemic; and, therefore, cannot be defined as infectious. In our approach, they would be included in *R*(*t*). This thought experiment explains why *I*(*t*) and *R*(*t*) should be dependent upon *P*_*c*_(*t*).

In a second thought experiment, imagine that a person has just infected a new person. In this case, the infecting person cannot reinfect that newly infected person nor can the newly infected person infect their infector. For the purposes of our model, we assume that *P*_*c*_(*t*), does not change quickly when compared to the rate that people become infected. This means that, mathematically, the infecting people durably remain in infectious contact with the people they have infected. Therefore, according to our definition, the infectiousness of the infecting people diminishes with each infection they cause; and, in symmetry, their recovery, the degree to which they are unable to infect others, increases.

In the third thought experiment, we recognize that during the course of an infection, the quantity and quality of the infectious agent within any infected person, the so-called “viral load”, will rise and fall with time. The ability of a person to infect susceptible people will also vary in synchrony, therefore, we postulate that the value of the infectiousness will approximately track the variation in viral load. It is easy to see that persons whose infection has passed the maximum viral load, will, in a static social situation, become increasingly less infectious; and, in a sense, partially recovered. Therefore, mathematically, infected persons, while clearly no longer susceptible, should be considered to be a part of both subpopulations *I*(*t*) and *R*(*t*).

Our intent in presenting these three examples is to provide intuitive insight into the characteristics we should expect for the variables *I*(*t*) and *R*(*t*) within the KMES. Specifically, the key characteristic is that *I*(*t*) is a measure of infectiousness which depends upon a person’s ability and, importantly, opportunity to infect others. Adopting this definition of *I*(*t*) leads inexorably to the conclusion that an individual will necessarily sometimes be a member of both the *I*(*t*) and *R*(*t*) subpopulations.

### Kermack and McKendrick’s model structure

Kermack and McKendrick (1927) derived their integro-differential equations by imagining *N*(*t*) to be the sum of incremental subpopulations of people who had been infected at the time *t* − *θ*; where *θ* was a dummy variable for time used to indicate the time since infection of each subpopulation. The time history of the epidemic was then imagined as a square t by t array with every successive row representing an increment of time, Δ*t*, and each column representing an increment of *θ*. We designate each increment of *θ* as Δ*t* since *θ* has the units of time.

The progress of each *θ* group in Kermack and McKendrick’s analysis was then tracked through time diagonally across the array with each *θ* group starting at time *t* − *θ* and progressing diagonally thereafter. The array is square because *θ* ≤ *t*; and Δ*θ* = Δ*t*. Their equations were then developed by initially summing over *θ* and then taking the limit as Δ*θ* and Δ*t* → 0. A full rendition of the arrays relevant to their equations is in Supplement 6. A close review of these arrays may be helpful for the reader to fully understand the relationship between time and *θ*.

To derive the solution to Kermack and McKendrick’s equation, we initially adopt their convention using *θ* and use the following notation. First, unless otherwise specified, all dependent variables and parameters are assumed to be both dependent on time, designated as t, and *θ*, defined as the time since infection. Examples of notation used to show this explicitly are *I*(*t*, *θ*) or *Ψ*(*t*, *θ*). If a variable (not a parameter) is designated as a function of time alone, then it is to be understood that the variable has been integrated over all *θ*. For instance, 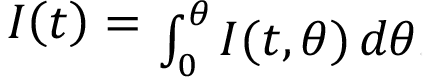. In contrast to the variables, however, if a parameter is designated as solely a function of time or *θ* alone, then the parameter is considered to be constant over the alternate temporal dimension; over either *θ* or time respectively. The dependent variables used in this manuscript are S, I, N, R; and the parameters are: *K*_*T*_, *P*_*c*_, φ, *φ*_*r*_, ρ, ψ, µ, β, *P*_*cr*_, and *γ*.

We also note that Kermack and McKendrick, in their manuscript, showed the parameters, φ, ψ, and β, at times as solely dependent on either *θ* or t, which is sometimes confusing. For clarity, we choose to initially show these parameters as dependent on both *θ* and t (i.e., we write them as φ(*t*, *θ*), ψ(t, *θ*), and β(*t*, *θ*)) and then let the analysis determine whether they are solely dependent on *θ*, t, or both. As the analysis proceeds, the importance of this clarification will become apparent.

With the prior concepts of *K*_*T*_(*t*), *P*_*c*_(*t*), *I*(*t*) and *R*(*t*) in mind, we write the Kermack and McKendrick integro-differential equations in terms of time and *θ*:

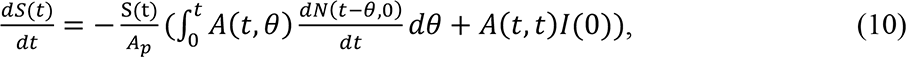

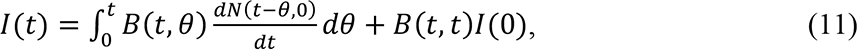

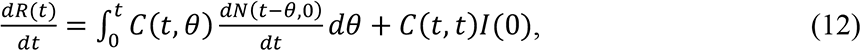

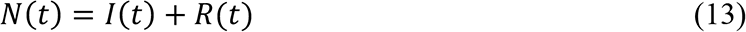

where 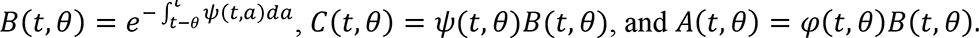.

Kermack and McKendrick (1927, p. 703) defined *φ*(*t*, *θ*) as “the rate of infectivity at age *θ*”, and *Ψ*(*t*, *θ*) as “the rate of removal” (page 703) of the infected population to the recovered population. *A*_3_ is the area that contains the population and, as previously defined, *θ* is the time since infection of any member of the population *N*(*t*).

The integration limits in the integral in the definition of *B*(*t*, *θ*) require some additional explanatory details. In Kermack and McKendrick’s development of their equations, they assumed that the *B* term, the infectiousness of each *θ* group at time, t, was only a function of *θ* because they assumed that infectiousness only represented the progress of the infections within each *θ* group. They chose, therefore, to assume that every *θ* group would proceed along identical time paths from the time since infection, *θ*. Consequently, the integration limits in the *B* term were chosen to only depend upon the time since infection (i.e., 0 to *θ*), regardless of the actual interval. They should have been more precise in their notation. The appropriate limits are *t* − *θ* and *t* because this is the actual time period associated with each *θ* group.

It was the recognition of these integration limits that enabled the solution which is demonstrated later in this section. We surmise that this subtle imprecision in Kermack and McKendrick’s notation; and the misinterpretation of the units of the parameters are likely the reasons that subsequent authors have not successfully found a pathway to a solution to the integro-differential equations.

### Derivation of the KMES

Our goal is to find a solution to Equations 10-13 in terms of t, *K*_*T*_(*t*) and *P*_*c*_(*t*). To develop this solution, we first convert Equations 10-13 into equations dependent on the variable t alone using *K*_*T*_(*t*) and the parameter µ(*t*). We then solve these new equations, express the solution in terms of *K*_*T*_(*t*) and *P*_*c*_(*t*); and use the resulting equations to develop expressions for *B*(*t*, *θ*), *φ*(*t*, *θ*), and *Ψ*(*t*, *θ*) to complete the solution. Lastly, we make the simplifying assumption that the epidemic parameters, *K*_*T*_(*t*) and *P*_*c*_(*t*) are constants, which leads to a simple set of intuitive expressions describing the epidemic dynamics.

We begin by using the definition of *N*(*t*) as the total number of people that have been infected. Therefore:

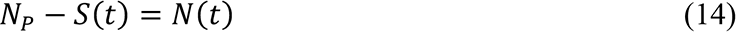

and,

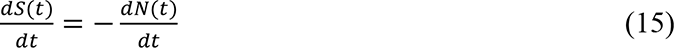

We have already defined *K*_*T*_(*t*) in Equation 1 as the rate of new infections caused by *I*(*t*), therefore,

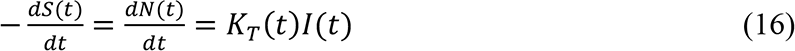

Equation 16 describes a direct relationship between the cause of the epidemic, namely, infections, *I*(*t*), and the change in the susceptible population; therefore, the units of *K*_*T*_(*t*) must 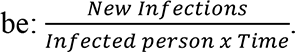

If we divide Equation 10 by Equation 11, and combine with Equation 16, we can express *K*_*T*_(*t*) in terms of Kermack and McKendrick’s variables as the negative of the change in the susceptible population per infectiousness, *I*(*t*):

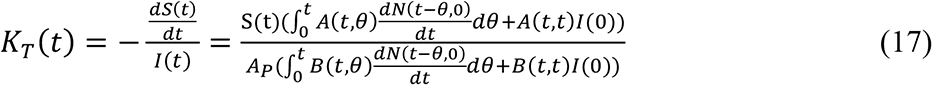

Likewise, by dividing Equation 12 by Equation 11 we can also define the parameter, µ(*t*), as the change in the recoveries per infectiousness, *I*(*t*):

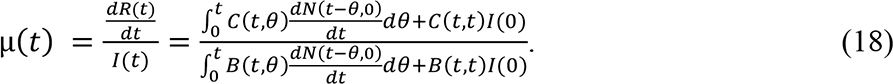

We note here that, as defined, µ(*t*), is only a function of time, meaning that for all values of *θ* at a given time, t, µ(*t*) has the same value. The same is true for the parameter *K*_*T*_(*t*).

We then rewrite Equations 10-13 in the following form as functions of time,

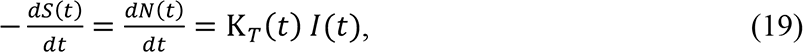

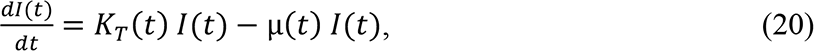

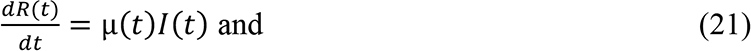

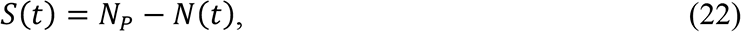

where *N*(*t*) = *I*(*t*) + *R*(*t*). We now use Equations 10 and 11 along with Equations 19 and 20 to find a relationship between µ(*t*) and *Ψ*(*t*, *θ*); and then a relationship between *K*_*T*_(*t*) and *φ*(*t*, *θ*).

Equation 20, is, of course, identical to Equation 2, and its solution is therefore Equation 3 from the introduction,

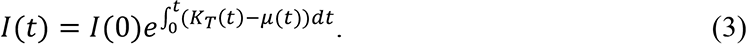

We now note that 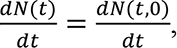, since the value of 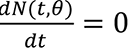 for all *θ* > 0. Then, since we know the relationship between 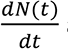 and *I*(*t*) from Equation 16, we can utilize the exponential forms of *B*(*t*, *θ*) and *B*(*t*, *t*), substitute Equations 3 and 16 into Equation 11, keeping in mind that *dθ* = *dt*, and find the following relationship,

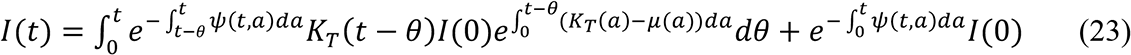

If we then divide Equation 23 by Equation 3, we find,

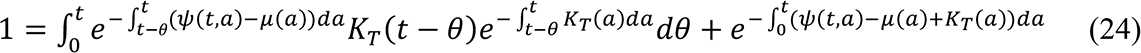

Since the integral, 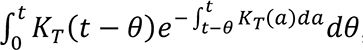, can be evaluated by using the relationship, 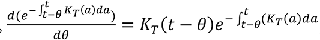, Equation 24 is satisfied if *Ψ*(*t*, *θ*) = *μ*(*t*), which is the first sought after relationship.

The second relationship, between *K*_*T*_(*t*) and *φ*(*t*, *θ*), can be found by substituting Equations 3 and 16 into Equation 10, using the exponential forms of *B*(*t*, *θ*) and *B*(*t*, *t*), and obtaining the following,

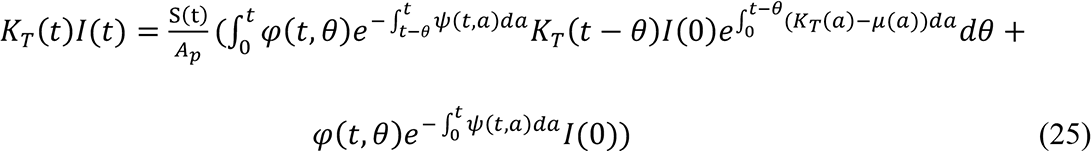

By using the relationship, *Ψ*(*t*, *θ*) = *μ*(*t*); dividing Equation 25 by Equation 3 and by *K*_*T*_(*t*), we find the following expression,

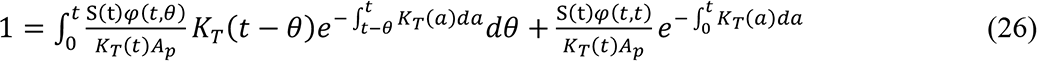

We then find that, as before, since 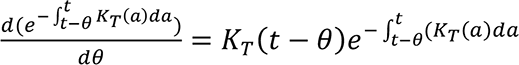, Equation 26 can be satisfied if 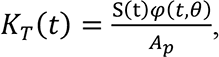, which is the second sought after relationship.

We now need to express µ(*t*) (and, therefore, *Ψ*(*t*, *θ*)) in terms of the epidemic parameters, *K*_*T*_(*t*) and *P*_*c*_(*t*). We begin by first dividing Equation 16 by *N*(*t*), integrating, and exponentiating to arrive at the following expression,

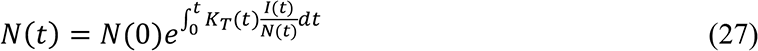

The remaining task is to find a relationship between 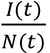 in terms of the parameters of the epidemic, *K*_*T*_(*t*) and *P*_*c*_(*t*) which we will then use to find the form of µ(*t*).

To this end, we continue the analysis by noting that every person within both *N*(*t*) and *I*(*t*) contacts *P*_*c*_(*t*) people in each period Δ*t*. Beginning at *t* = 0, then, according to Equation 1, a total of *K*_*T*_(0)*I*(0)Δ*t* people within the *I*(0)*P*_*c*_(0) group becomes infected during the time Δ*t*; and the number of non-infected people within the *I*(0)*P*_*c*_(0) group is changed to, *I*(0)*P*_*c*_(0) −*K*_*T*_(0)*I*(0)Δ*t* + *I*(0)Δ*P*_*cni*_(0); where *P*_*cni*_(*t*) is the number of people within *P*_*c*_(*t*) who are still susceptible. We add the term *I*(0)Δ*P*_*cni*_(0) to account for any additional susceptible people that the *I*(*t*) group may contact infectiously during the time interval Δ*t*. This also means that, per potentially infectious contact, *I*(0)*P*_*c*_(0), the infectiousness of the people within *N*(0), has been changed by the factor, 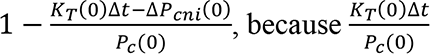 is the portion of *P*_*c*_(0) the contacting infected people can no longer infect and 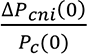 is the number of new contacts that can be infected.

After a time Δ*t*, the infectiousness, *I*(Δ*t*), in the affected population *N*(Δ*t*) is, therefore, per contact,

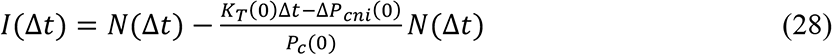

Dividing both sides by *N*(Δ*t*), we obtain a difference equation for the ratio of the infectiousness, *I*(Δ*t*), to the total of the ever infected, *N*(Δ*t*), population,

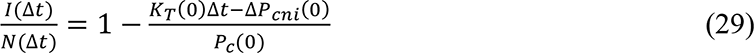

In the next time step, another *K*_*T*_(Δ*t*)Δ*tI*(Δ*t*) people within the group *P*_*c*_(Δ*t*) become infected and, therefore, the infectiousness within *N*(2Δ*t*) is changed by an additional factor, 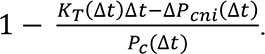. This is expressed mathematically as,

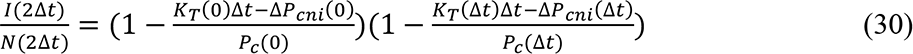

The process repeats itself in each period Δ*t*; and therefore, we can write the recursion formula,

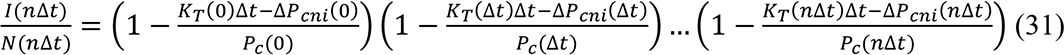

As we indicated in the second thought experiment, we now make the simplifying assumption that a change, Δ*P*_*cni*_(*t*), to *P*_*cni*_(*t*) will occur much more slowly than the creation of new infections, which means that Δ*P*_*cni*_(*n*Δ*t*) ≪ *K*_*T*_(*n*Δ*t*)Δ*t*. Therefore, in Equation 31, we can neglect the Δ*P*_*cni*_(*n*Δ*t*) terms. The situation where Δ*P*_*cni*_(*n*Δ*t*) is plausibly growing as fast as *K*_*T*_(*n*Δ*t*)Δ*t* corresponds to an outbreak from the main epidemic. This is explored in more detail in Supplement 5.1.

If we neglect the Δ*P*_*cni*_(*n*Δ*t*) terms in Equation 31; and, since by definition, *n*Δ*t* = *t*, as *n* → ∞, Δ*t* → 0, Equation 31 becomes,

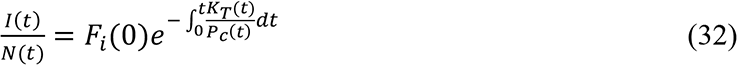

where 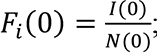; and is the fraction of *N*(*t*) that is infected at *t* = 0. Equation 32 is the sought for relationship for 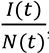, in terms of *K*_*T*_(*t*) and *P*_*c*_(*t*).

Substituting Equation 32 into Equation 27, we now have the following expression for

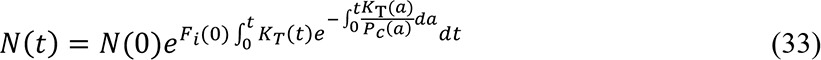

and, by multiplying Equations 32 and 33, we obtain an expression for *I*(*t*),

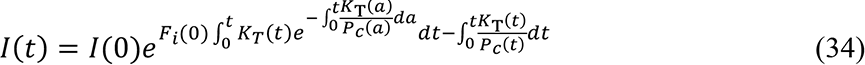

From the definition of *N*(*t*) and using Equations 33 and 34 we find the expression for

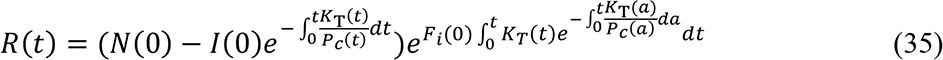

If we substitute Equations 33 to 35 into Equations 19 through 22, we find that they are a solution if 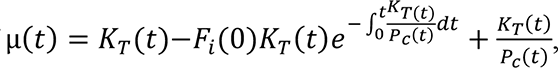, which is the sought after expression for µ(*t*).

To complete the solution to Kermack and McKendrick’s integro-differential equations, we use the expression for µ(*t*) to write the forms of *B*(*t*, *t*), *B*(*t*, *θ*), *φ*(*t*, *θ*), and *Ψ*(*t*, *θ*) in terms of *K*_*T*_(*t*) and *P*_*c*_(*t*),

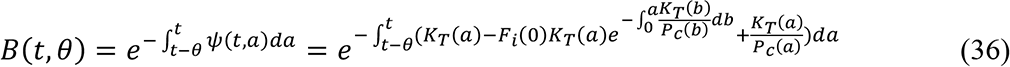

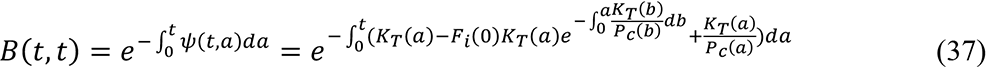

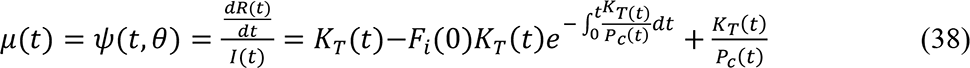

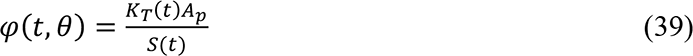

Equations 33 through 39 form a complete solution to Equations 10 through 13. We refer to this solution as the KMES (<u>K</u>ermack and <u>M</u>cKendrick <u>E</u>quation <u>S</u>olution) We also note that in Equations 38 and 39, *Ψ*(*t*, *θ*) and *φ*(*t*, *θ*) are only functions of time; and we therefore continue this manuscript with only the use of *Ψ*(*t*) and *φ*(*t*). Lastly, with no loss of generality, since *S*(*t*) = *N*_!_ − *N*(*t*), rather than communicate the progress of the epidemic in terms of susceptibles, *S*(*t*), we will continue to express it in terms of the total cases, *N*(*t*), as in Equation 33.

### Simplified expressions

As we indicated at the beginning of the KMES derivation, the complex solution in Equations 33 through 39 can be written in simplified, more intuitive forms. The first of these is what we term the “Step Response” form, which is derived by using Equation 37 to rewrite Equations 33 through 35 as,

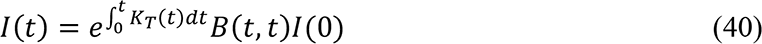

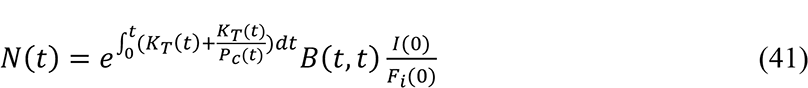

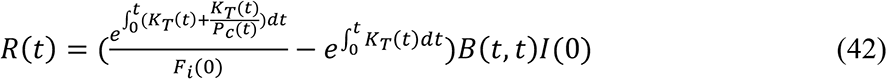

Since *B*(*t*, *t*) is the time varying infectiousness input of the original infected group, *I*(0), the exponential expressions, 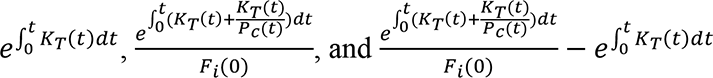, are the step response functions to this input. Equation 40 also shows that if there were no recovery, that is, if *B*(*t*, *t*)*I*(0) = 1, then the epidemic would proceed exponentially until the entire population was infected, an intuitive and sensible result.

In a final simplification of the solution, as an approximation, if we assume that both *K*_*T*_(*t*) and *P*_*c*_(*t*) are constant for a period in Equations 33 through 35 and if *F*_*i*_(0) = 1, we arrive at simplified statements for *N*(*t*), *I*(*t*), and *R*(*t*),

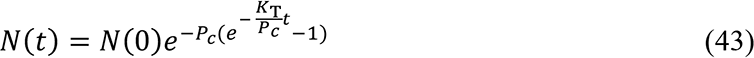

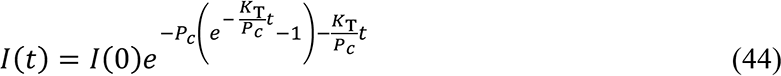

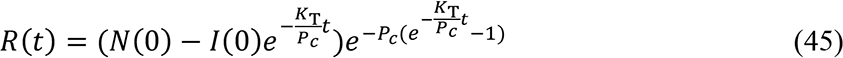

## Section 2: Useful expressions derived from the solution

In this section we illustrate the many additional useful expressions that can be derived from the KMES. For simplicity, we assume that *K*_*T*_ and *P*_*c*_ are constants and that *I*(0) = *N*(0) = 1 = *F*_*i*_(0). These simplifying assumptions allow the nature of the expressions to be more easily seen and understood.

### Herd Immunity

If *K*_*T*_ truly is a function of the disease alone, it is likely to be a constant, at least in the initial stages of an epidemic. Using this inferred property, and assuming the population does not change its behavior (i.e., *P*_*c*_ is a constant) during the pandemic, we find the following expression for the final size, *N*(∞) as *t* → ∞, using Equation 43,

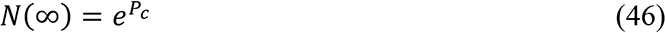

From Equation 46, we can see that, if *P*_*c*_ is large enough, it is possible for *N*(∞) to equal *S*(0) and, therefore, herd immunity is not guaranteed.

The time it would take for *N*(∞) = *S*(0) is given by the following expression,

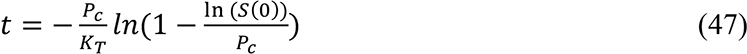

and the criteria that must be true for the entire population to become infected is,

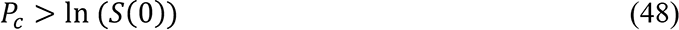

Equations 46 through 48 enable the estimation of the level of social interaction that will cause the total population to be infected and they demonstrate very directly that the existence of herd immunity is not an inherent property of the Kermack and McKendrick model.

### Basic and Effective Reproduction Number, R0 and REff

If we divide *K*_*T*_ by *Ψ*(*t*) we find an expression for the Effective Reproduction Number (*R_Eff_*):

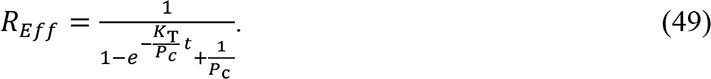

A function of both the disease and the behavior of the population, the value of *R*_*Eff*_ marks two key epidemic points. First, the peak in the number of new infections occurs and the epidemic begins to decline when *R*_*Eff*_ = 1. Second, when *t* = 0, *R*_*Eff*_ = *R*_0_ = *P*_*c*_ = *Basic Reproduction Number*.

### Time to Peak New Infections

By setting *R_Eff_* = 1 in Equation 49, we obtain the following expression for the time when the decline in new cases, 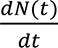 begins:

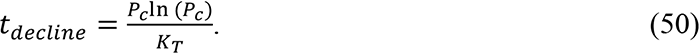

Likewise, if we differentiate both sides of Equation 43 twice, we can obtain an identical expression for the time when 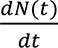 is a maximum,

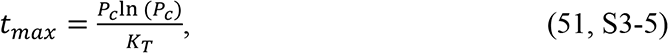

where *t_max_* = the time to the peak of new infections. As it should, the time of the peak in new cases coincides with the start of the decline of infections.

Equation 51 demonstrates the relationship between the strength of social intervention measures, *P*_Y_, and the time to the peak of new infections. When social interventions are stronger (smaller *P*_Y_), the time to the peak will always be shorter.

### Measuring the Control of an Epidemic

A key purpose in developing a closed form model is to use expressions such as Equations 46 to 51 to determine whether control measures in place are adequately managing the epidemic.

Here we develop tools from the KMES for assessing the condition of the epidemic in real time to enable adjustments to control measures.

First, by differentiating both sides of Equation 33, dividing both sides by *N*(*t*), rearranging the terms, and then taking the natural log of both sides, we obtain the following useful expression,

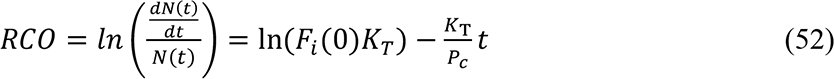

We label this the “Rate of Change Operator” (RCO) because it is a measure of the rate of change of *N*(*t*), per person within *N*(*t*). The RCO is a convenient expression to use in evaluating the current epidemic conditions because it only requires knowledge of the recent data on total number of cases, *N*(*t*), and daily new cases, 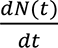, and as we show later, estimates of both *K*_*T*_ and *P*_*c*_ can be made from the value of the RCO.

Another important expression, derived by differentiating Equation 43 twice, is the rate of acceleration of the epidemic:

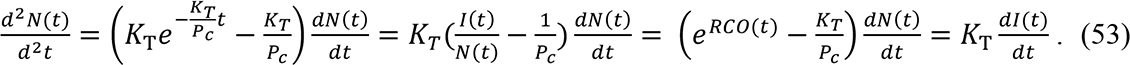

Equation 53, with its four equivalent expressions, demonstrates the power that an authentic model provides. The leftmost expression allows us to compare the acceleration—the potential to change the rate of new infections—at any stage of the epidemic for any two countries, even those with different population densities, using only the daily case rate and the defining constants, *K_T_*_’_ and *P_c_*.

Equation 53 is also an immediate determinant of whether the control measures in place, represented by *P*, are effective enough. If the value of the term, 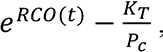, in the third equality is positive, then the control measures are not strong enough. Conversely, when this term is negative, the epidemic is being brought under control.

The maximum value of *P*_*c*_ that will begin to bring down the new cases per day is the value that turns the acceleration in Equation 53 negative. If we set the left-hand side of Equation 53 to zero, use the third expression from the left in Equation 53 and solve for *P*_*c*_, we arrive at the defining relationship for this critical objective of epidemic management:

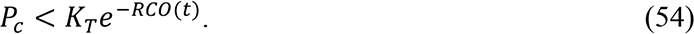

Since the value of *RCO*(*t*) can be easily determined every day by calculating 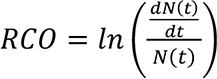 during the epidemic and the value of *K*_*T*_ can be determined using the technique illustrated in Section 3, the maximum allowable value of *P*_*c*_ needed to reduce the number of daily cases can always be determined. This value of *P*_*c*_ is the maximum level of infectable social contact allowable if we want the number of new daily cases to continue decreasing. Also, as explained in Supplement 5.1, if the slope of the RCO curve is determined from the graphical analysis to be greater than zero, then an outbreak has occurred and immediate reductions in social interactions are needed.

If we define a desired target for 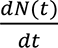, the number of new cases per day, at a future time, *t* + *t_target_*, we can derive a new quantity, the desired fraction of the current new cases, *D_tf_*, as:

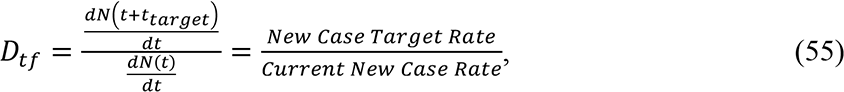

Using the derivative of Equation 43, we arrive at the following expression:

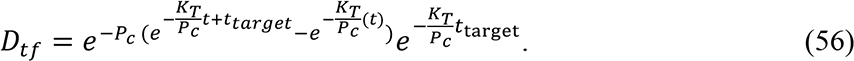

If *t* ≫ *t_target_*, then 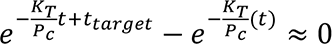 and we obtain the following equation from the remaining terms:

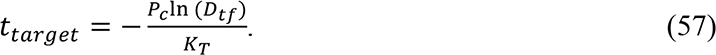

Equation 57 quantitates the number of days, *t*_%@A\8%_, that a level of social containment, *P*_*c*_, must be imposed to achieve a fraction of daily cases, *D*_%;_, compared to the current level. In Supplement 5, using quantitative examples, we explain the use of Equations 53 through 57 to characterize and control an epidemic.

## Section 3: Veracity of the solution

We can test the KMES projections using data from the Covid-19 pandemic. To do this, we must first determine the appropriate values of *K*_*T*_ and *P*_*c*_.

### Projecting Country Data

If we assume that both *K*_*T*_ and *P*_*c*_ are at least piece-wise constant for substantial periods of time, we see that Equation 52 is a linear equation in time. By applying Equation 52 to data (Roser, et al. 2021) from six different countries during the initial stages of the Covid-19 pandemic, we obtained the curves in Figure 1. As can be seen in the figure, before and shortly after the date (indicated by the arrows in the figures) of the imposition of containment actions in the six countries, these RCO curves became straight lines. This is a strong indication that it is reasonable to assume that *K*_*T*_ and *P*_*c*_ were approximately constants both before and after the imposition of the containment actions.

**Figure 1.**
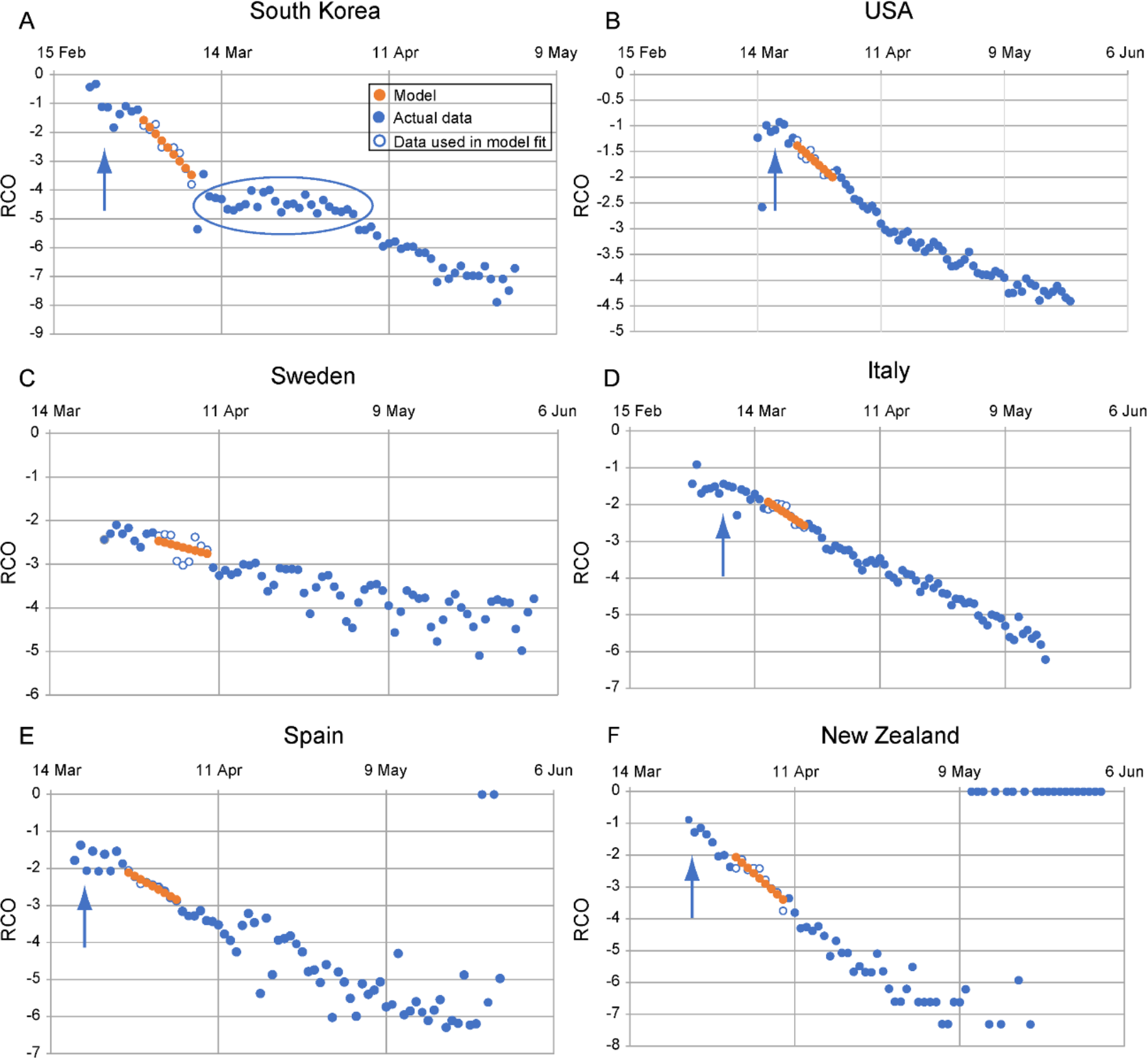
Rate of change operator (RCO) curves for COVID-19 cases in various countries. An epidemic can be described by a piecewise linear model using the RCO (Equation 52). A short segment of orange dots in each graph is a linear fit to the corresponding points (blue/white circles) in the observed data. The slopes and initial points of these dotted-line segments are the values of *K*_1_ and *K*_2_ respectively which are tabulated in Table1. In some countries, RCO curves changed markedly soon after the date containment measures were implemented (arrows): **A)** South Korea, February 21; (the oval highlights a departure of the observed data from the RCO slope, indicating failures in, or relaxations of, social distancing); **B)** USA, March 16; **C)** Sweden did not implement any specific containment measures, so the model calibration was begun on April 1, the date when the slope of the RCO curve first became steady. **D)** Italy, March 8; **E)** Spain, March 14; **F)** New Zealand, March 25. All dates are in 2020.

To project the course of the epidemic in the individual countries, we used a small portion of the data immediately after the imposition of the containment measures to determine the values of ln(*F*_*i*_(0)*K*_*T*_) and 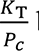 by fitting Equation 52 to short, early portions of the straight segments of the RCO time series. These early portions comprised nine data points each and Table 1 displays the values derived for each country.

**Table 1.** Social containment parameters used to model total cases and new daily cases of infection for different countries (Roser et al 2021).

| | $\frac{K_T}{P_c}$ | $\ln(F_i(0)K_T)$ | $N_{(0)}$ | Date range for RCO fit |
| --- | --- | --- | --- | --- |
| **South Korea** | 0.24 | -1.58 | 3,736 | March 1–March 9 |
| **USA** | 0.076 | -1.39 | 46,136 | March 23–March 31 |
| **Sweden** | 0.036 | -2.47 | 5,320 | April 1–April 9 |
| **Italy** | 0.080 | -1.93 | 31,506 | March 17–March 25 |
| **Spain** | 0.09 | -2.11 | 65,719 | March 27–April 4 |
| **New Zealand** | 0.17 | -2.06 | 708 | April 1–April 9 |

Using these values in Equation 43, we then predicted the course of daily total cases (Figure 2) for the six countries. These predictions matched the actual time series of the daily <u>total</u> cases with an *R^2^* > 0.97 in each of the six countries for the 45 days following the date containment measures were introduced. We then used the derivative of Equation 43 to plot the predicted time series of the daily <u>new</u> cases in Figure 3 for the six countries for the same 45 days.

**Figure 2.**
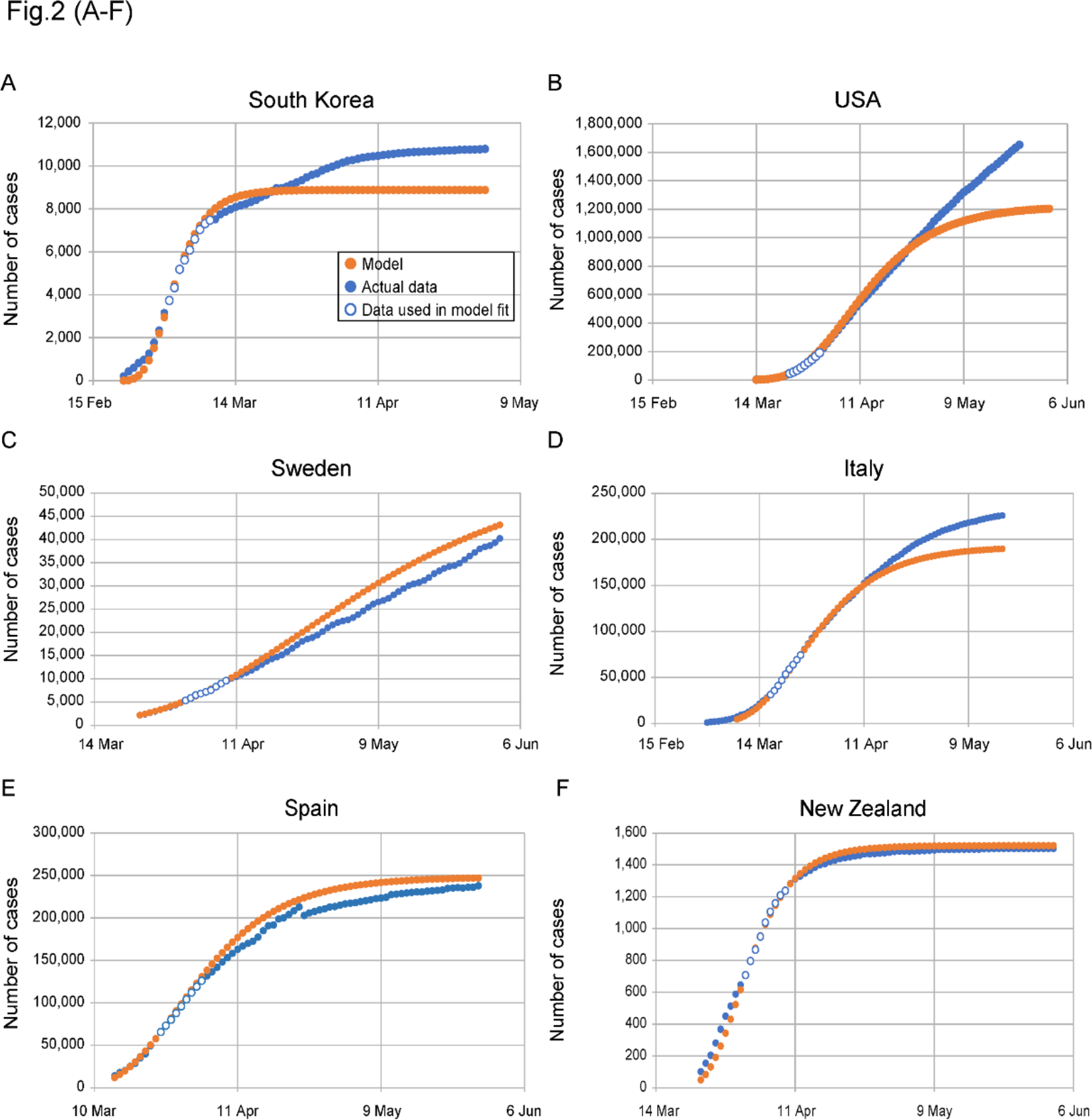
Complete KMES model predictions for daily total case counts. **A)** South Korea; **B)** USA; **C)** Sweden; **D)** Italy; **E)** Spain; and **F)** New Zealand. Dots are daily data points observed from (white-center and all blue) or calculated (orange) for each country. The KMES model was calibrated using data from the date ranges listed in Table 1 (white-center blue dots). *R*^2^ > 0.97 for the model fit for all countries for the 45 days after the containment measures were implemented: South Korea, February 21-April 4; USA, March 16-April 30; Italy, March 8–April 22; Spain, March 14-April 28; New Zealand, March 25-May 9. Sweden did not implement any specific containment measures, so the dates used were March 23- May 7. The deviation of the model from the data in the USA, panel (**B**), after April is elucidated in Supplement 5. All dates are in 2020.

**Figure 3.**
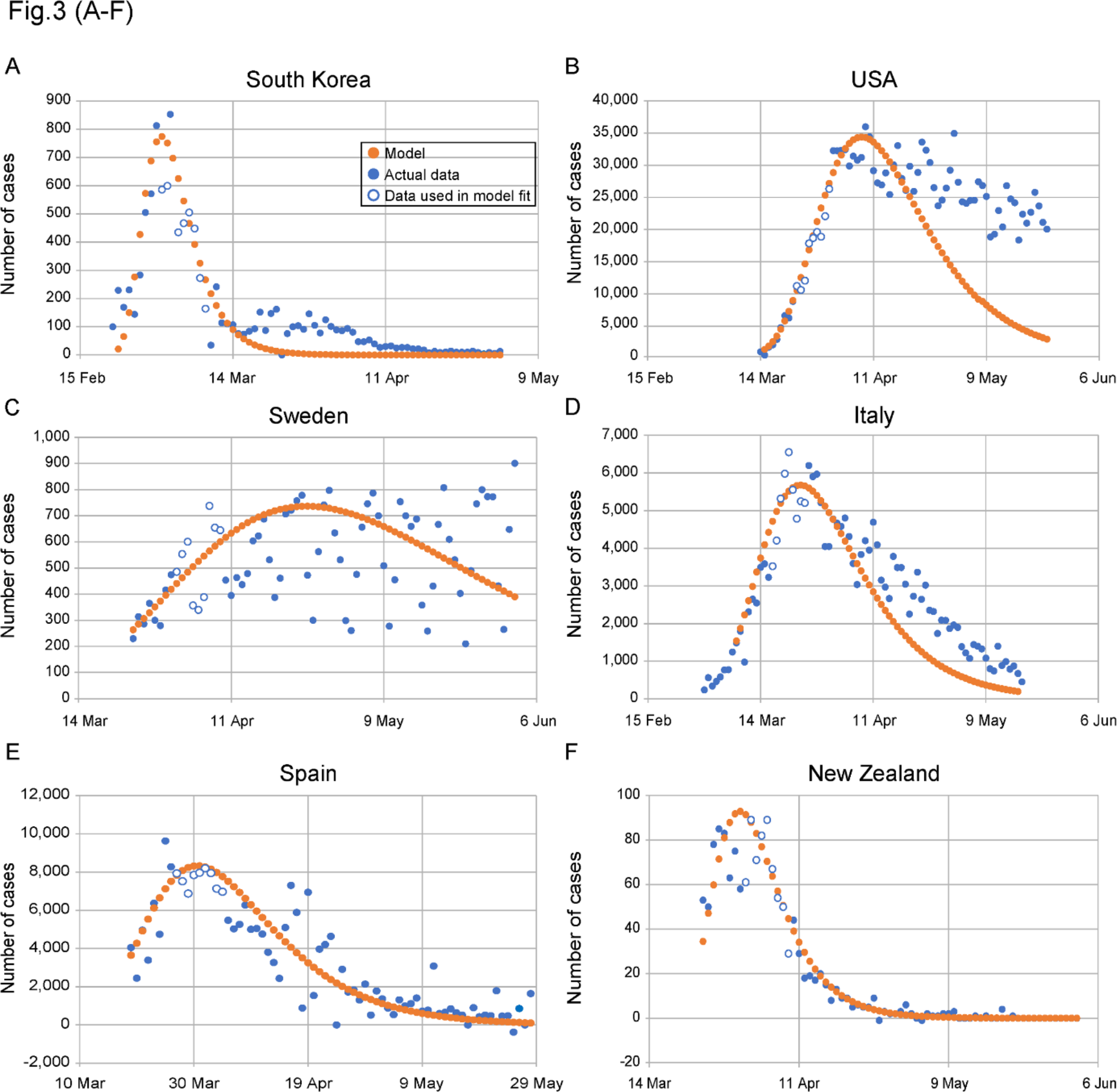
Complete KMES model predictions for number of new daily cases. **A)** South Korea, *R*^2^ = 0.86; **B)** USA, *R*^2^ = 0.83; **C)** Sweden, *R*^2^ = 0.29; **D)** Italy, *R*^2^ = 0.69; **E)** Spain, *R*^2^ = 0.65; and **F)** New Zealand, *R*^2^ = 0.90. The orange dotted line is the model in all panels. The all-blue and white-center blue dots are data points, daily observations from each country. The white-center blue points are used to determine model parameters. *R*^2^ values are between the model and the data, across countries for the 45 days after containment measures were instituted.

These predictions have an *R^2^* range of 0.29 to 0.90; and as seen in the figure; the predicted peak of new cases was close to the observed peak for all countries.

It is important to emphasize that the predictions in Figures 2 and 3 are *not* fits to the full-length of the data shown. Rather, the two constants, ln(*F*_*i*_(0)*K*_*T*_) and <u>^*8^</u> were estimated using only a short, linear, nine-point portion of the epidemic data starting between 7 to 14 days after the imposition of containment measures. These constants were then used to project the data for the days after the nine points.

### *K*_*T*_(*t*) as a Parameter of the Disease

In an additional demonstration of the veracity of the KMES, we tested the assumption that, in the initial portion of the epidemic, *K*_*T*_(*t*) is a constant and a property of the disease, and therefore, should be the same for each country. Equation 58, S2-5, derived in Supplement 2, shows that the model parameters, expressed in a purposefully constructed function,

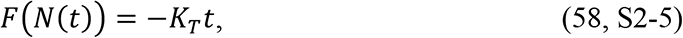

Where 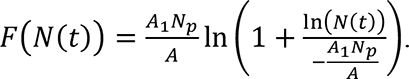 should be linearly proportional to time with a constant of proportionality or slope equal to −*K*_*T*_. As illustrated in Figure 4, the fit of Equation 58, using the population density data from Table 2, has an *R^2^* = 0.956 and a slope of −0.26 (the slope is equal to −*K*_*T*_). This excellent correlation confirms that *K*_*T*_ can confidently be assumed to be the same for all countries, is a constant in the initial stage of the epidemic, and is likely, a property of the disease.

**Figure 4.**
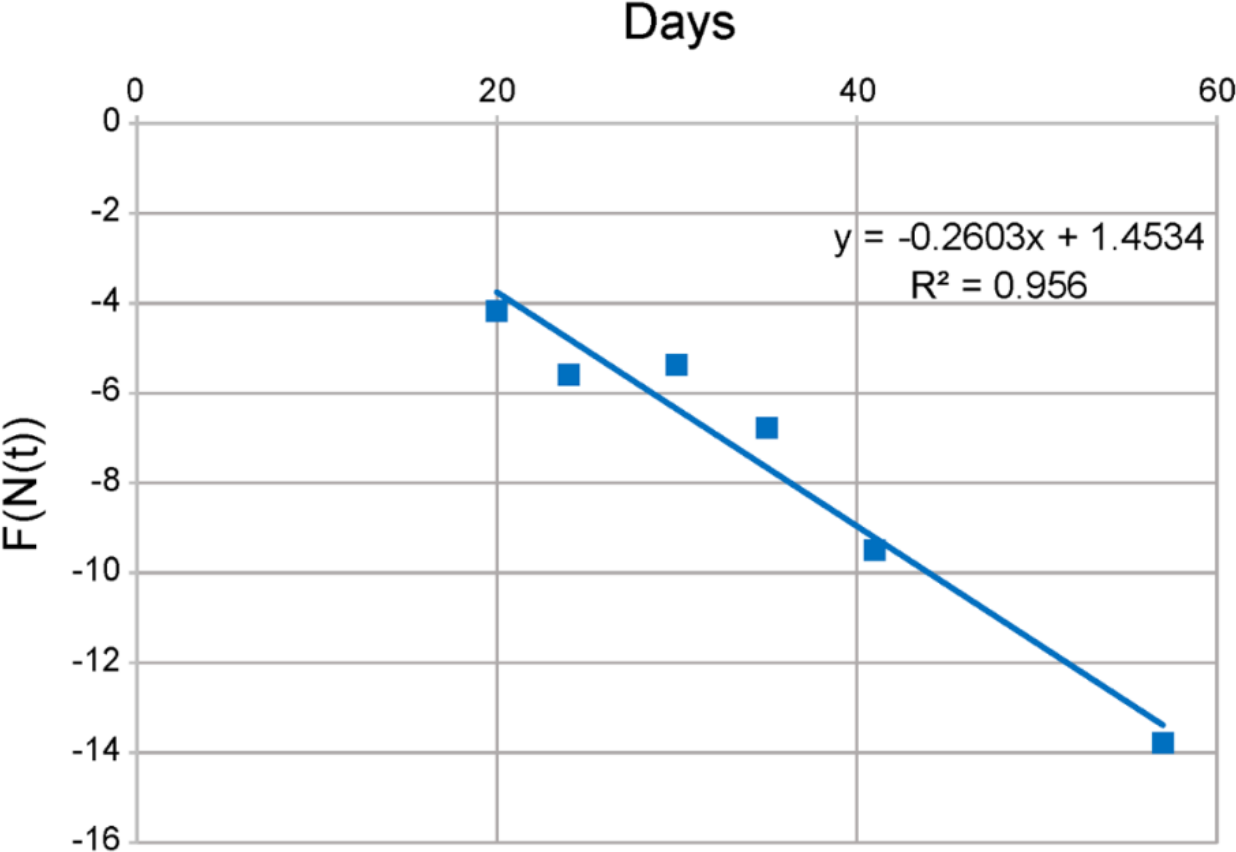
Verification that *K*_*T*_ is the same for all countries. The data from Table 2 is plotted using Equation 58 and *A*_H_ = 0.48 *km*^2^. Each data point corresponds to a different country. The value of *K*_*T*_ is the negative of the slope of the line, and *K*_*T*_ is closely approximated everywhere by *K*_*T*_ ≈ 0.26

**Table 2.** Initial COVID-19 pandemic data and social interaction parameters for various countries ((Roser et al 2021), case and date data; (Worldometers 2021, population density data)

| | Date of first case reported | Date of cases in calculation | Days | Cases on calculation date | Population density (people/km<sup>2</sup>) | $K_T$ | $\ln(K_T)$ | $\frac{K_T(0)}{P_c(0)}$ |
| --- | --- | --- | --- | --- | --- | --- | --- | --- |
| **South Korea** | 22 Jan | 21 Feb | 30 | 204 | 527 | 0.26 | -1.3 | 2.39E-04 |
| **USA** | 22 Jan | 19 Mar | 57 | 13,663 | 36 | 0.26 | -1.3 | 3.50E-03 |
| **Sweden** | 1 Feb | 7 Mar | 35 | 179 | 25 | 0.26 | -1.3 | 5.04E-03 |
| **Italy** | 31 Jan | 24 Feb | 24 | 229 | 206 | 0.26 | -1.3 | 6.11E-04 |
| **Spain** | 1 Feb | 13 Mar | 41 | 5,232 | 94 | 0.26 | -1.3 | 1.34E-03 |
| **New Zealand** |  | 19 Mar | 20 | 28 | 18 | 0.26 | -1.3 | 6.86E-03 |

All dates are in 2020.

### Viral Load

The step response structure of the KMES suggests that it should also contain an expression for the viral load of the disease. Equation 37 provides a starting point because it describes the evolution of the infectiousness of the initially infected population. If we set *P*_*c*_ and *I*(0) = 1 in Equation 37, meaning that if there was only one initial infection and we assume that this infected person contacted, on average, only one person during the epidemic, and assume for this case, that *K*_*T*_(*t*) is a constant, we obtain an expression for the infectiousness of the initially infected person,

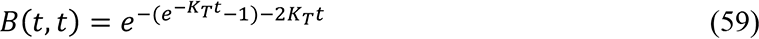

However, as we have noted, infectiousness does not equate to the actual viral load.

Rather, it is a measure of the ability to infect and is dependent on the portion of contacts that remain infectable as well as the viral load. Since the available portion of infectable people in *P*_*c*_ is equal to *R*_*Eff*_, we can extract the viral load from the infectiousness by dividing *B*(*t*, *t*) by *R*_*Eff*_ (dividing Equation 59 by Equation 49), which results in the following expression (keeping *P*_*c*_ = 1),

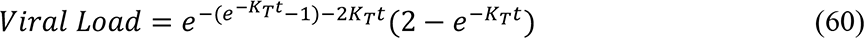

When we set *K_T_*= 0.26, as derived from Figure 4, we obtain the plot in Figure 5.

**Figure 5.**
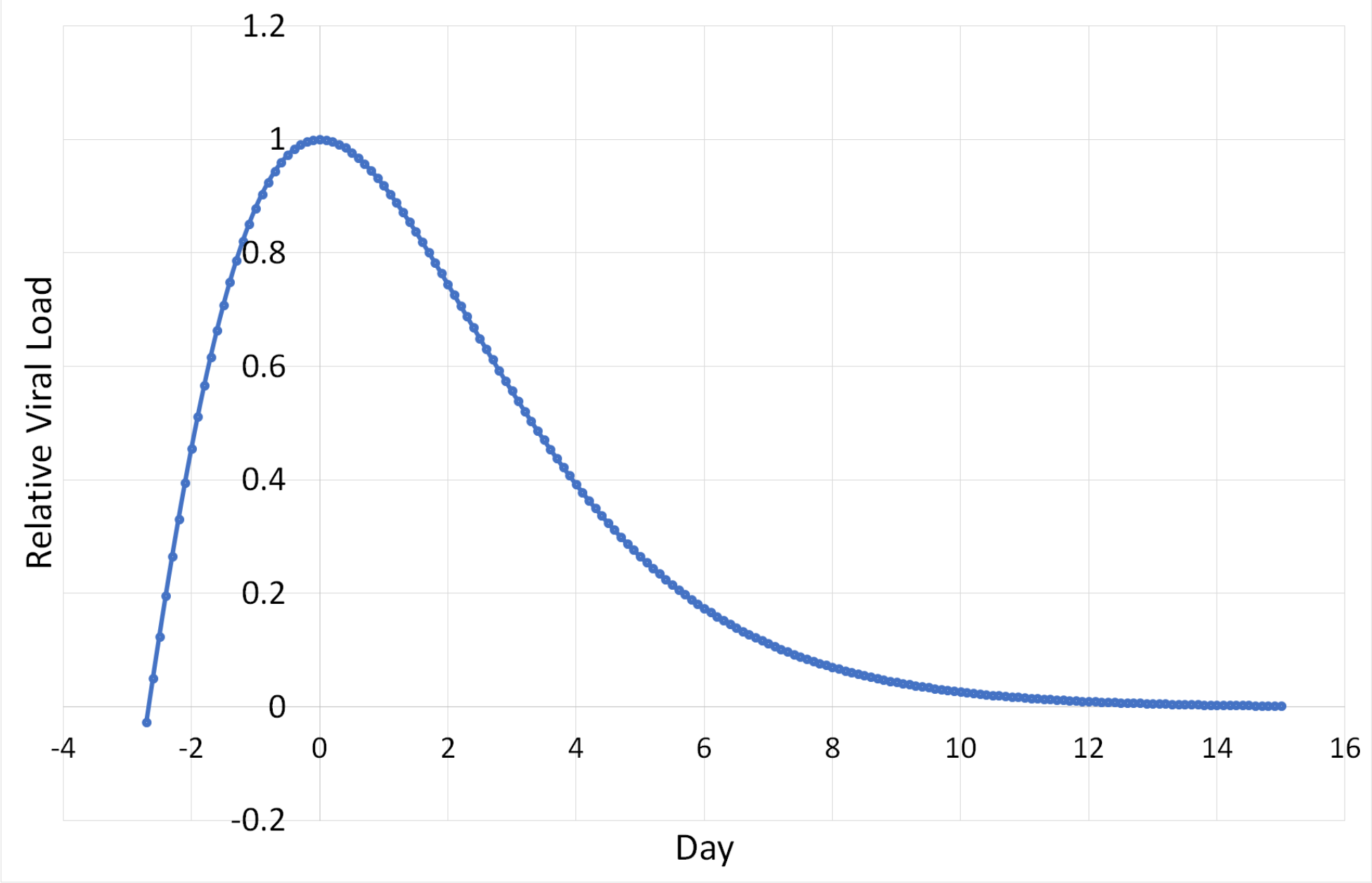
Representation of the average viral load of an infected person in the Covid 19 pandemic. The plot was generated using the value *K*_*T*_ = 0.26 in Equation 60

The interpretation of Equation 60 and its graphical representation in Figure 5 require careful consideration. First, the initially infectious person in the epidemic did not become infected at *t* = 0. Therefore, time in Equation 60 and in Figure 5 must be allowed to be negative at the beginning of the infection. Second, Equation 60, as seen graphically in Figure 5, has the expected characteristics of a viral load; that is, it rises quickly, reaches a maximum, and then declines at a slower exponential pace.

As a measure of the veracity of the curve in Figure 5, we found that it has the same overall shape and dynamic change in load from peak to 15 days after the peak as the estimated average viral loads estimated by direct measurements of individual Covid-19 patients with Covid-19 (Challenger et al 2021 and Jones et al 2022). In these references, using patient data, Challenger et al (2021) found that the base 10 exponential decline rate of the viral load was -0.22 with a credible interval between -0.26 and -0.17 whereas Jones et al (2022) found a credible interval of the base 10 exponential decline of -0.17 to -0.16. A least squares base 10 exponential curve fit to the data in Figure 5 between day 0 and day 15 has a rate of decline of -0.19 with an R^2^ value of 0.99. This result is another strong indication of the veracity of the KMES and that *K*_*T*_ is a parameter of the disease.

### Correlation with Independently Measured Mobility Data

A fourth illustration of veracity arises from the ability to correlate independently sourced Google mobility data (Google 2020) to the RCO. Mobility data, available from Google, are a measure of the difference between the amount of time people stayed at home (the Residential Mobility Measure or RMM) during the period modelled and a baseline measured for 5 weeks starting January 3, 2020. As explained in Supplement 2, and based on the expression for the RCO (Equation 52), if the KMES is correct, then the integral of this mobility data should correlate linearly with the measured RCO. Figure 6 shows that, as the KMES predicts, for each country considered, the integral of the RMM and the RCO are linearly correlated to a high degree.

**Figure 6.**
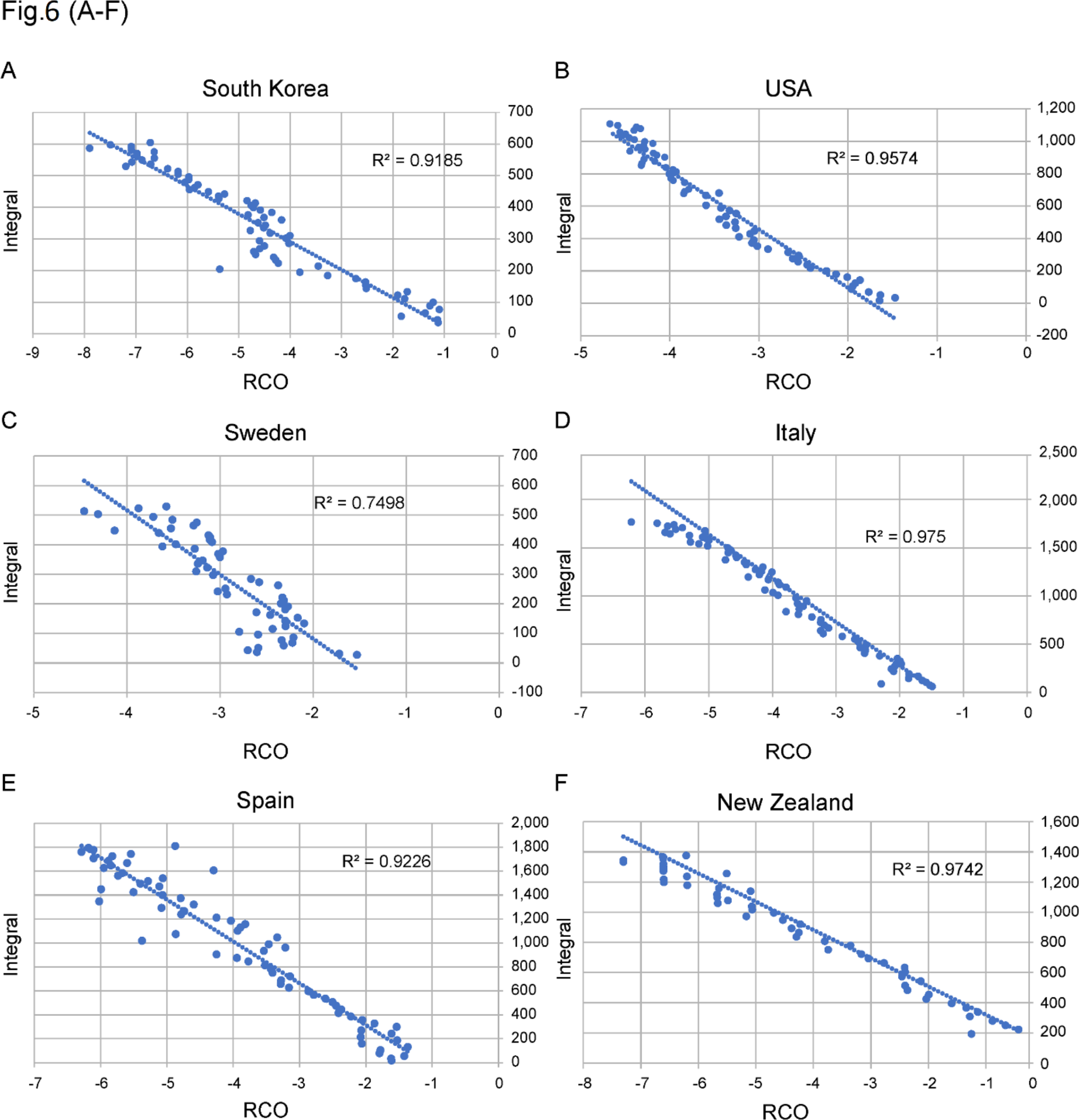
Correlations between the daily value of the rate of change operator (RCO) and the integral of the Google Residential Mobility Measure (RMM) (Google 2020). **A)** South Korea (date range, February 23 to April 23); **B)** USA (March 25 to May 31); **C)** Sweden (March 5 to May 5); **D)** Italy (March 25 to May 31); **E)** Spain (March 25 to May 31); and **F)** New Zealand (March 21 to April 22). All dates are in 2020.

## Section 4: Comments on the SIR approximations

Due to the previous lack of a closed form solution, approximations to the full Kermack and McKendrick integro-differential equations appear throughout the literature. Since, by their definition, approximations differ from the exact form, it is instructive to analyze these approximations with an eye to determining whether they behave even qualitatively like the KMES solution; and whether conclusions based on the approximations are then likely to be valid.

The approximations have their roots in Kermack and McKendrick’s 1927 paper where they proposed the following approximation,

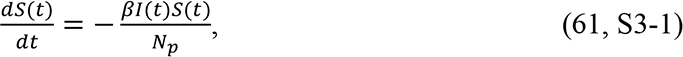

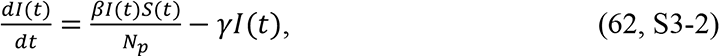

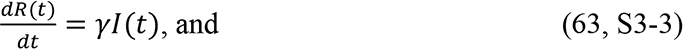

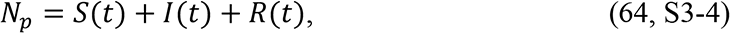

where *β* = rate of contact and transmission, *γ* = rate of recoveries. It should also be noted that the basic reproduction number 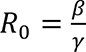 by definition.

These well-known “SIR” equations can be derived from the full Kermack and McKendrick equations by assuming that the parameters 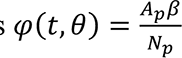 and *Ψ*(*t*, *θ*) = *γ* are constants. The SIR equations and their variants (SEIR, MSEIR, etc.) have been used for decades in attempts to quantitatively and qualitatively model epidemics. These models are known as compartmental models, and they all share the common characteristic that the term 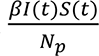 appears in the equations for 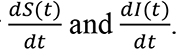

In Section 1 we derived a time-based form of the Kermack and McKendrick equations; Equations 19 through 22. Although Equations 61 through 64 are purported to be an approximation to these equations, we find that if *K*_*T*_(*t*) and µ(*t*) in Equations 19 through 22 are approximated as the constants β and *γ* as in the SIR model, the two sets of equations are not even approximately equivalent because the term 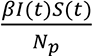 does not appear in the approximations of Equations 19 or 20. Since the SIR equations are putatively an approximation of the <u>full</u> Kermack and McKendrick integro-differential equations and Equations 19 through 22 are just a restatement of Kermack and McKendrick’s equations; this suggests a potential flaw in the methodology used to derive the SIR equations. Therefore, we need to re-examine the logic Kermack and McKendrick used in developing the SIR equations. In the following subsections, we focus in particular on the reasons why *φ*(*t*, *θ*) and *Ψ*(*t*, *θ*) should not be approximated as constants.

### Why φ should not be approximated as a constant

The argument against approximating *φ* as a constant originates with a key statement put forth early in the derivation of Kermack and McKendrick’s integro-differential equations. On page 703 Kermack and McKendrick state (1927), “Now v(*t*) denotes the number of persons in unit area who become infected at the interval *t*, and this must be equal to 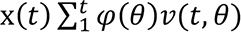 where x(*t*) denotes the people per unit area still unaffected (the density of the susceptibles), and *φ*(*θ*) is the rate of infectivity at age *θ*.” In equation form, using our notation of *φ*(*t*, *θ*), this statement reads,

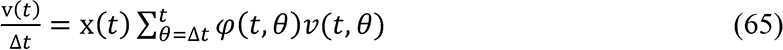

Later in their manuscript, Kermack and McKendrick used Equation 65 as the basis for deriving Equations 60 and 61 by assuming that *φ*(*t*, *θ*) equals the constant 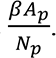.

Although Kermack and McKendrick labeled *φ*(*t*, *θ*) as a “rate”; a proper balancing of the dimensions in Equation 65 demonstrates that *φ*(*t*, *θ*) cannot merely be a rate with the units of *time*^-1^. Because v(*t*, *θ*) and v(*t*) have the same units, 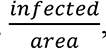, and *X*(*t*) has the units of 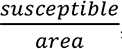, we must conclude that *φ*(*t*, *θ*) has the units of 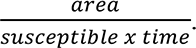. Hence, from the description given, it is clear that *φ*(*t*, *θ*) should be defined in the following form using the actual rate,

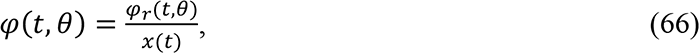

where *φ*_*r*_(*t*, *θ*) is a rate of new infections. Using Equation 66, Equation 65 can now be rewritten as,

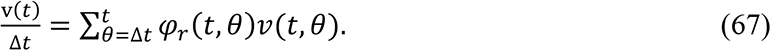

In an epidemic, as the number of total infections increases, the density of the susceptibles, *x*(*t*), is an ever-decreasing quantity; and therefore, from Equation 66 we can see that *φ*(*t*, *θ*) cannot itself be considered a constant as the epidemic progresses; especially as the affected population grows very large. Rather, the quantity, *φ*_*r*_(*t*, *θ*) (which is equivalent to *K*_*T*_(*t*)) is a better choice to be considered a constant when seeking to simplify the equations without losing their essence.

Other authors too, have misinterpreted and misused *φ*(*t*, *θ*). For example, in Hethcote (2000, pg 602), the term, 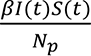 is derived using the argument that “If *β* is the average number of adequate contacts (i.e., contacts sufficient for transmission) of a person per unit time, then 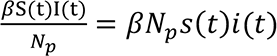 is the number of new cases per unit time due to *S*(*t*) = *N*_*p*_*s*(*t*) susceptibles.” Notable in the statement is that *β* is defined as, “…the average number of adequate contacts (i.e., contacts sufficient for transmission) of a person per unit time …”. Because *β* is defined as the average number of adequate contacts per unit time, ***<u>and per person</u>***, this statement also implies that *β* is not merely a contact rate with the units of 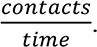. Since Heathcote goes on to imply that the “person” is a susceptible, *β* must have the units 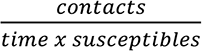 and the conclusions that 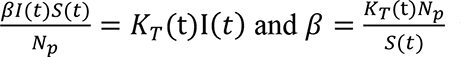 immediately follow.

Early in an epidemic, when *S*(*t*) ≈ *N*_3_, we see that *φ*(*t*, *θ*) ≈ *φ*_*r*_(*t*, *θ*) = *K*_*T*_(*t*). Therefore, *φ*(*t*, *θ*) can only be approximated as a constant early in an epidemic and if *K*_*T*_(*t*) can be approximated as a constant at that point.

### Why Ψ should not be approximated as a constant

The time early in an epidemic, when *S*(*t*) ≈ *N*_3_, also illustrates why *Ψ* should not be approximated as the constant γ in the SIR formulation. When *S*(*t*) ≈ *N*_3_, Equations 60 through 63 become,

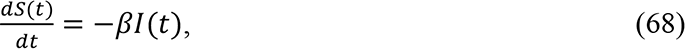

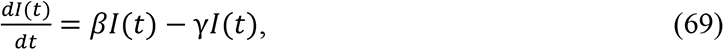

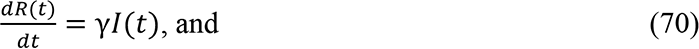

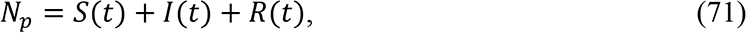

Since peaks are observed in epidemics even when *S*(*t*) ≈ *N*_3_ and *β* = *φ*(*t*, *θ*) ≈ *K*_*T*_(*t*) early in an epidemic; and, as explained earlier, *K*_*T*_ is a parameter of the disease which is likely to be a constant early on, inspection of Equation 69 makes it immediately clear that this set of equations will not adequately model epidemics. Equation 69 predicts a continual exponential increase in *I*(*t*) at a constant rate of *β* − γ if *β* > γ. Therefore, in contradiction to observed phenomenon, Equation 69 predicts a peak will never occur while *S*(*t*) ≈ *N*_3_. Since *β* can be approximated as a constant under these conditions, we are forced to conclude that *Ψ* cannot be adequately modelled as a constant while *S*(*t*) ≈ *N*_3_. This conclusion is supported by Equation 38 which shows that ψ(*t*), and therefore *γ*, cannot be a constant because the term, 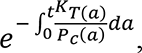 decays exponentially with time.

From the preceding observations and since we have the KMES, we conclude that all SIR constructs are inappropriate and unnecessary approximations. In Supplement 3, we support this conclusion by detailing the mathematically implausible assumptions that underlie the SIR approximations and make the case that these assumptions are not even qualitatively correct. We also analyze the inherent flaws within the SIR formulation that led to the erroneous “flatten-the-curve” projection by explicitly demonstrating that the flatten-the-curve projection illustrated throughout the literature (see, for example, Di Lauro et al 2021) is caused by hidden, inherent and implausible assumptions in the SIR models about both the populace and the disease.

## Discussion

The derivation of a solution to the Kermack and McKendrick integro-differential equations obviates the need for an approximation to these same equations. Rather than use an approximation, such as the SIR models, we can instead use the solutions for *N*(*t*), *I*(*t*), and

*R*(*t*), illustrated in Equations 33 through 39, to predict the dynamics of an epidemic, determine whether the societal controls in place are adequately managing the epidemic, and develop quantitative measures for guiding the behavior of the populace, all goals of epidemic management. We also illustrate that the phenomena predicted using the SIR approximation are not, in fact, properties of the Kermack and McKendrick equations.

We began this manuscript with three observations:

1. A fundamental tenet of modern epidemiology, the presumed existence of herd immunity, is premised on a circular argument;
2. The phenomenon of the peak in new cases moving out in time with declining values of *β* in the SIR model, actually stems from less efficient transmission among a highly interactive population, and;
3. The observation that the peak in infections in the SIR model precedes the peak in new cases is an indication that unknown assumptions about the population behavior lie hidden within the SIR model.

Using the KMES we fully explicate the mathematical bridges between the KMES and the SIR models and demonstrate why the SIR approximation is a poor substitute for a closed form solution to the full integro-differential equations.

The KMES accurately projects phenomena which arose in the Covid epidemic, even under the simplifying assumption that *K*_*T*_(*t*) and *P*_*c*_(*t*) are constants for periods of time. These successful projections contrast sharply with SIR model projections which must violate the assumption that *β* is constant to produce even approximately consonant results. Also, the high degree of correlation between the proxy of the population behavior in the KMES and the independently measured actual population mobility in the Google data contrasts with the weak correlations of the SIR construct to mobile phone mobility data found by prior authors (Wesolowski 2015). These accurate projections and the contrasts with the SIR approximations, strongly suggest that the KMES should replace the SIR models.

Our expression for final epidemic size, an important finding, disproves another of the accepted tenets of epidemiology. Equations 46 through 48, mathematical expressions for that final size, the time to reach it and the associated contact required, in their mathematical simplicity, state what is an intuitive conclusion: if the people contact each other infectiously at a high enough level for a long enough time, herd immunity is not guaranteed, and an epidemic <u>can</u> spread to an entire population. This conclusion must become a new tenet of epidemiology.

The KMES does have the intuitive form, exemplified by the systems view encapsulated in Equation 40:

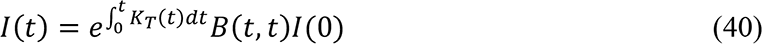

This equation states that the input of infections, *B*(*t*, *t*)*I*(0), is transformed into the time varying output of infectiousness, *I*(*t*), through an exponential step response function, 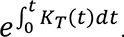. Our analysis here has led us to affirm this practical mathematical statement of the epidemic dynamics.

As further support to the veracity of our solution, we note that expressions for the number of total cases, *N*(*t*), Equations 29 and 42, are both Gompertz equations. This form is supported by Onishi et al (2021) who demonstrate that the Covid-19 epidemic time course in many countries was well fit by a Gompertz model. These authors do not offer a basic principles argument as to why this is so, but they demonstrate a strong correlation to this aspect of our model structure.

With the availability of an analytical solution, we derived previously unknown, pragmatically useful expressions of important epidemiological relationships: Time course of the epidemic size, Final epidemic size, Time to peak infections, Effective Reproduction number, Viral load, and targets for reducing the epidemic along a planned path. In addition, in the supplements, we explain how to detect and model outbreaks. These relationships form an important new toolbox for public health officials to use in accurately guiding the public to control an epidemic.

These analytical expressions are intuitive and sensible. For instance, the expression for time to maximum new cases, in Equations 50 or 51, passes smoothly through the epidemic peak. This contrasts with the expression for the time to the peak derived from the SIR models (Koger and Schlickeiser, 2020). In this reference, the expressions for the SIR models are only valid when *R*_*Eff*_ > 1. While we do not doubt that they have been correctly derived from the SIR equations, they do show that the SIR model has the peculiar property that the estimated time to the peak in new infections becomes increasingly larger as *R*_*Eff*_ approaches one before suddenly plunging to negative infinity just as *R*_*Eff*_ reaches one. This mathematically summarizes the claimed phenomena behind the concept of “flattening the curve”, but it is unsettling and nonintuitive. How can the peak in new infections actually move away from attainment as people interact less frequently?

Our expressions for the time to the peak, Equations 50 and 51, have none of this peculiar behavior and their forms are supported by data from different countries which imposed very different containment strategies. As we explain in Supplement 3, when social containment is increased, the peak number of infections is much lower, and it occurs earlier. The KMES shows that strong containment actions shorten the epidemic, as one would intuit; and the data from several countries clearly demonstrate. This finding is also supported by the data presented in Figure 4 in Harris (2023).

The fundamental reason conventional SIR models project epidemic phenomenon incorrectly is that beginning with Kermack and McKendrick, numerous authors have misunderstood the units of *φ* on the way to deriving the SIR equations. Even authors (Hethcote 2000), who derived the SIR equations without the use of Kermack and McKendrick’s equations, replicate this mistake. Furthermore, *Ψ*(*t*) cannot be assumed constant early in the epidemic; and *φ*(*t*) cannot be assumed constant as the epidemic becomes large relative to the total population without imposing subtle, implausible assumptions on the model. Analytic expressions for these quantities, Equations S3-8 and S3-9, derived in Supplement 3, make this clear.

It should not be surprising that a closed form, complete solution to the epidemic equations produces an expression for the viral load. The curve in Figure 5, wholly derived from the Covid-19 data from several countries, has the characteristics many authors have expected a viral load to have (Challenger et al 2022, Jones et al 2021). While these authors reached their conclusions through direct measurement of the viral load of thousands of patients, the same form has emerged from the KMES by only using country case data.

Equations 34 and 35 clearly show that both *I*(*t*) and *R*(*t*) depend on both *K*_*T*_(*t*), a property of the disease and *P*_O_(*t*), a function of the population behavior. It is natural to assume that both *I*(*t*) and *R*(*t*) will depend on properties of the disease, but the form of the KMES shows directly that their values also depend on the behavior of the population.

We explain this dependency in Section 1 and Supplement 1 by showing that *I*(*t*) is best interpreted as the total infectiousness within the infected population *N*(*t*). As a complementary interpretation, *R*(*t*) is best thought of as the degree of recovery from infectiousness within *N*(*t*). Therefore, a previously infected individual is simultaneously a part of both the infected and recovered populations with the degree of membership determined by the parameter ψ(*t*).

As time goes on, the degree of membership inevitably moves the infected individuals towards membership in the recovered community, but during this time, the infectiousness of all individuals vary with their viral load and number of contacts. An increase in social contact causes an increase in infectiousness, which, in turn, decreases the degree to which the person remains in the recovered population and vice versa. Therefore, as an individual’s viral load changes, based on time and the disease dynamics, so too, does this individual’s ability to infect others change based on their level of social interaction.

With this concept of variable membership in mind, then, we see that the idea of a compartmental model wherein people move irreversibly from being infected to recovered is an inadequate model construct. Rather, assuming immunity exists, the proper compartment construct is that there are only two compartments: 1) not yet infected, S(*t*); and 2) previously infected, *N*(*t*); and only from the latter of these is there no escape.

## Concluding remarks

We recognize that the mathematics and resulting conclusions described in this manuscript contradict long-accepted, mathematically derived tenets of epidemiology. However, these conclusions have been derived by directly solving the integro-differential Kermack and McKendrick equations, the foundation of epidemic models. While the abandonment of long held concepts is always a difficult proposition, it is nevertheless necessary given the demonstrations here that prior approximations do not even qualitatively approximate important, real world phenomenon. The proposed replacement for these concepts, the KMES, is consistent with and provides insight into both data gathered from the Covid-19 pandemic and basic epidemiological notions such as the Effective Reproduction Number and viral load. The expressions derived using the KMES are well behaved under all epidemic conditions and accurately predict correlations among a variety of independent data sources.

We recognize that our analysis can be improved by an exploration of population interactions that differ from the ones we assumed. We made the simplifying assumption that the ratio 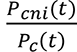 remains constant as *P* changes. This is a reasonable assumption, but an analysis which does not require its use may provide even deeper insights into epidemic behavior and management. The analysis can also be further improved by using the enormous amount of case data now available. This additional data can improve the estimate of the key parameter, *K*_*T*_(*t*), including variations in time (with mutations of the infectious agent) and possibly with local genetic variations in the population affected. This will further illuminate the actions people need to take to achieve the target values of *P*_*c*_(*t*).

We intend this work as a hopeful message to the epidemiological community. Logical, analytical tools are available to characterize the state of an epidemic and provide guidance to public health officials. These tools show, unequivocally, that with stronger initial measures, an epidemic can be stopped more quickly with much less economic damage than predicted by conventional models. Although each disease agent will have its own infectious process, the overall epidemic dynamics can ultimately be modified by the behavior of the population; and the KMES provides the detailed pathway to modification and perhaps, even control.

## Data Availability

All data produced in the present study are available upon reasonable request to the authors

## Supplement 1 Insights developed from the KMES

As we derived the KMES, we did not stop to discuss insights provided by some of the important expressions. In this section, we will provide those insights.

The first of these expressions is the relationship described by the derivative of Equation

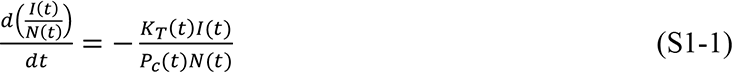

This seemingly simple expression is, along with Equation 1, a fundamental statement of an infectious epidemic.

If we write out the derivative, 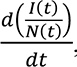, in Equation S1-1, we obtain:

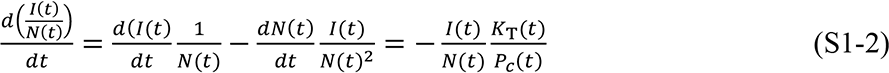

With some rearrangement of the terms, we find the following expression:

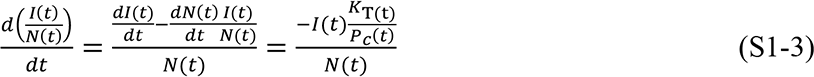

We highlight Equation S1-3, because it brings to light an important insight when we set *t* = 0, and *I*(0) = *N*(0). Setting *I*(0) = *N*(0) and *t* = 0 in Equation S1-3 and recognizing that 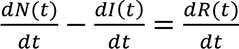 we obtain the following:

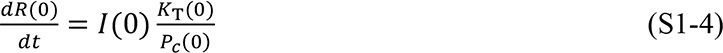

Since all three quantities on the righthand side are positive, Equation S1-4 provides a possibly startling result: The recovered population begins to grow the instant the epidemic starts!

Furthermore, Equation S1-4 tells us that the infectiousness of the individuals in the population *I*(0) (and by extension *I*(*t*)) is not a constant during the time they are infected. This may be obvious because, the viral load changes as the infectiousness of a person changes.

However, Equation S1-4 handily provides us with the initial rate at which the infectiousness of population *I*(0) is changing. That rate is 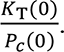

It is not surprising that disease infectiousness is a function of *K*_*T*_ but it is perhaps less obvious that the infectiousness changes directly and inversely to the population behavior, *P*_Y_.

The infectiousness of a person is not just dependent on the viral load, but also and comparably so on the contacts a person has with other, not-yet-infected people. Semantically, if an infected person never contacts another noninfected person, they are never truly infectious in the sense that they cannot advance the disease.

We can describe *R*(*t*) in an obverse manner, since *R*(*t*) is the reduction in the total infectiousness that has occurred in the population, *N*(*t*); and so, it is also a function of the disease transmissibility and population behavior.

We gain further insight into the meaning of the KMES by taking the derivative of Equation 44 and dividing by *I*(*t*). Then, using Equation 32 we obtain,

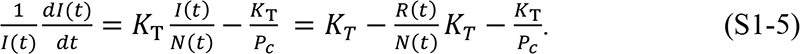

The left-hand side of Equation S1-5 is the rate of change of infections per infectious person. Since 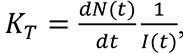, and is the rate at which infectious persons cause new infections, the terms 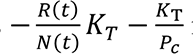 must be the rate of recovery per infectious person, 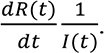

Finally, we can use Equations 33, and 46 to write this simple expression for the solution for total cases if *K*_*T*_(*t*) and *P*_*c*_(*t*) remain constant:

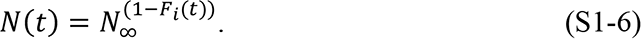

where 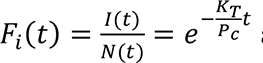 and is the fraction infected.

## Supplement 2. Verification of the solution

### Demonstration that *K*_*T*_ is a parameter of the disease and constant early in the epidemic

To demonstrate that *K*_*T*_(*t*) is indeed a constant, we need to first further refine the concept of *P*_*c*_(*t*). As previously defined, *P*_*c*_(*t*) is the average number of specific infectious contacts a member of subpopulation *N*(t) has across the entire population. This is obviously a function of the population’s behavior. Here, we begin by also assuming that *P*_*c*_(*t*) is a function of population density, and that people’s mobility extends over a constant average effective area per unit of time which we define as the effective area rate, *A*_SA_(*t*). We can then write an expression for *P*_*cr*_(*t*):

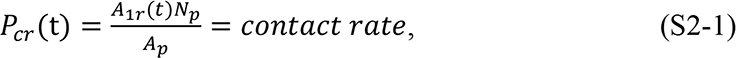

where *N*_3_ = the entire population of the region with the infection, = the area of the region, and 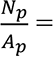 the population density.

Analogous to the way we defined *P*_*c*_(*t*) using *P*_*cr*_(*t*), we now define a quantity, *A*_S_(*t*), in terms of *A*_SA_(*t*):

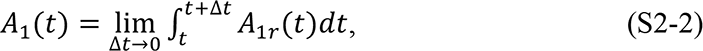

where *A*_S_(*t*) is the effective area traversed by an individual. In specifying the area, we make an assumption similar to the assumption that we made for *P*(*t*)_<_ in Section 1; that is, the area that a person traverses can change, but we assume that if a change in area changes the ratio of contacted persons not infected to total contacts 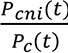, it does so slowly. We call *A* (*t*) the “effective area” because the population is typically only dispersed within ∼1% of the land within a given region of a country (Ritchie and Roser 2019). If we take this into account, then *A*_S_(*t*) = 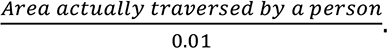. As defined, neither *P* (*t*) nor *A* (*t*) are rates. They are both unitless, vary in time, and depend on the population’s behavior.

We can now write an expression for 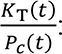

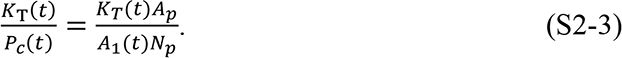

and we can check the assumption that *K*_*T*_(*t*) is a constant by substituting Equation S2-3 into Equation 33, 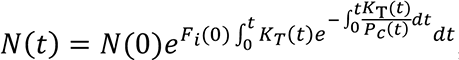, assuming that both *N*(0) and *F*_*i*_(0) are equal to 1; assuming both *K*_*T*_(*t*) and *P*_*c*_(*t*) are constants, and solving for *K*_*T*_t. Doing this, we find the following expression:

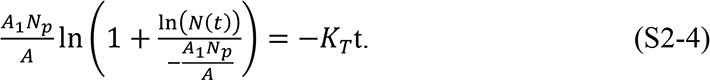

If we define 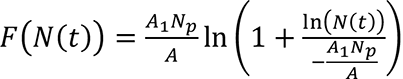, then we can also write this expression as

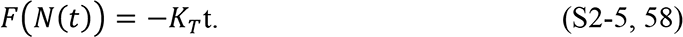

If *K*_*T*_ is a constant, then Equation S2-5 predicts that *FN*(*t*)| is a linear function of time.

Excepting *A*_S_, all the quantities on the left-hand side of Equation S2-4 can be found for each country in the time before containment measures were enacted; and these are listed for the sample of countries addressed in this paper, in Table 2. An implicit assumption in this process is that the behavior of the population, *P*_*c*_(*t*), is constant and therefore *A*_S_is constant at least during the initial phase of the epidemic before containment measures were put in place.

Because of this linkage, it is necessary to frame the problem as co-determining a value of *A*_S_ which produces a straight line for the country data; and separately determining whether the slope of that line is a rational value for *K*_*T*_. Through a process of iteration, a value was found for *A*_S_(= 0.48 km^2^) which created a straight line with a correlation coefficient of 0.96 (Figure 4). Using Equation S2-4 (or S2-5) we then determined the slope of the line in Figure 4 indicating the value of *K*_*T*_ as 0.26. This value is completely consistent with the country data; and we take this analysis as strong support for the plausibility that *K*_*T*_ is a constant across countries and represents the transmissibility of the disease in the early stage of the epidemic.

### Demonstration that *K*_*T*_ is a measure of population behavior

Independent evidence that *P*_*c*_(*t*) is a measure of the population behavior was developed by first using Equation 52 to show that, if *K*_*T*_(*t*) is a constant, the RCO measure will be proportional to 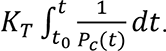. We reasoned that if an independent measure of people’s mobility during the epidemic could be identified and this was linearly related to the RCO, we could have additional confidence in the veracity of the KMES.

Google has compiled different measures, derived from mobile phone data, of people’s mobility (Google 2020). One of these measures is termed the Residential Mobility Measure (RMM). The RMM is a measure of the percentage change in the degree to which people stayed in their residence during the pandemic relative to a baseline measured over 5 weeks starting on January 3, 2020. Since 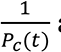 and the RMM are both inversely proportional to the population’s mobility, we hypothesized that the RMM might be a good proxy for the value of 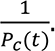. To test this, we plotted the integral over time of the daily RMM for the six countries whose data we analyzed, against the daily RCO. These plots appear in Figure 6; and they clearly support the hypothesized linear relationship.

## Supplement 3. An analysis of the SIR model

The SIR model, with constant parameters *β*and *γ*, is described by the following equations:

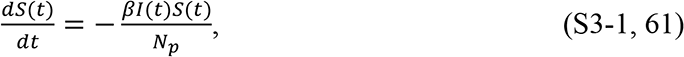

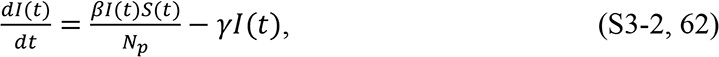

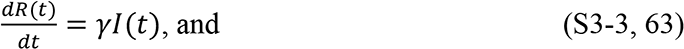

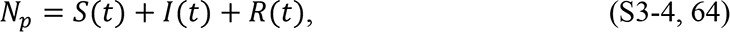

where *Np* = total number of people in the population, *β* = rate of contact and transmission, and *γ* = rate of recoveries. These equations can be derived from Equations 10 through 13 by assuming that the parameters 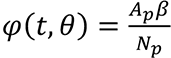 are constants.

### 3.1 “Flattening the Curve”

Since 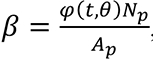, and *φ*(*t*, *θ*) was defined by Kermack and McKendrick (1927) as “the rate of infectivity at age *θ*” (page 703), *β* has generally been interpreted as an inverse measure of social containment in the at-risk population, i.e., modelers have assumed that a lower *β* indicates higher social containment. Likewise, since *γ* = *Ψ*(*t*, *θ*) and Kermack and McKendrick defined *Ψ*(*t*, *θ*) as “the rate of removal” (page 703) of infected persons to a recovered state or death, *γ* is generally interpreted as a measure of persistence of infectiousness, a constant associated with the agent of the disease; a lower *γ* has been assumed to represent longer-lasting disease.

A simulation created using an Euler approximation of Equations S3-1 through S3-4, depicted in Figures 7A and B, shows that the SIR model projects an increase in social containment (decreasing *β*) causes a later end to the epidemic and a lower and progressively later peak in cases per day. This is the so-called “Flatten the Curve” phenomenon predicted by Equations S3-1 through S3-4 which is often referenced in the literature (see Di Lauro, et al, 2021, as an example). In contrast, a plot of the KMES in Figures 7C and D exhibits the opposite phenomenology: an increase in social containment (higher <u>^*!^</u>) causes an *earlier* end to the epidemic and a lower and progressively *earlier* peak in cases per day. As social containment measures increase, the positions of the peak in new cases per day move in *opposite* directions for the two models.

**Figure 7.**
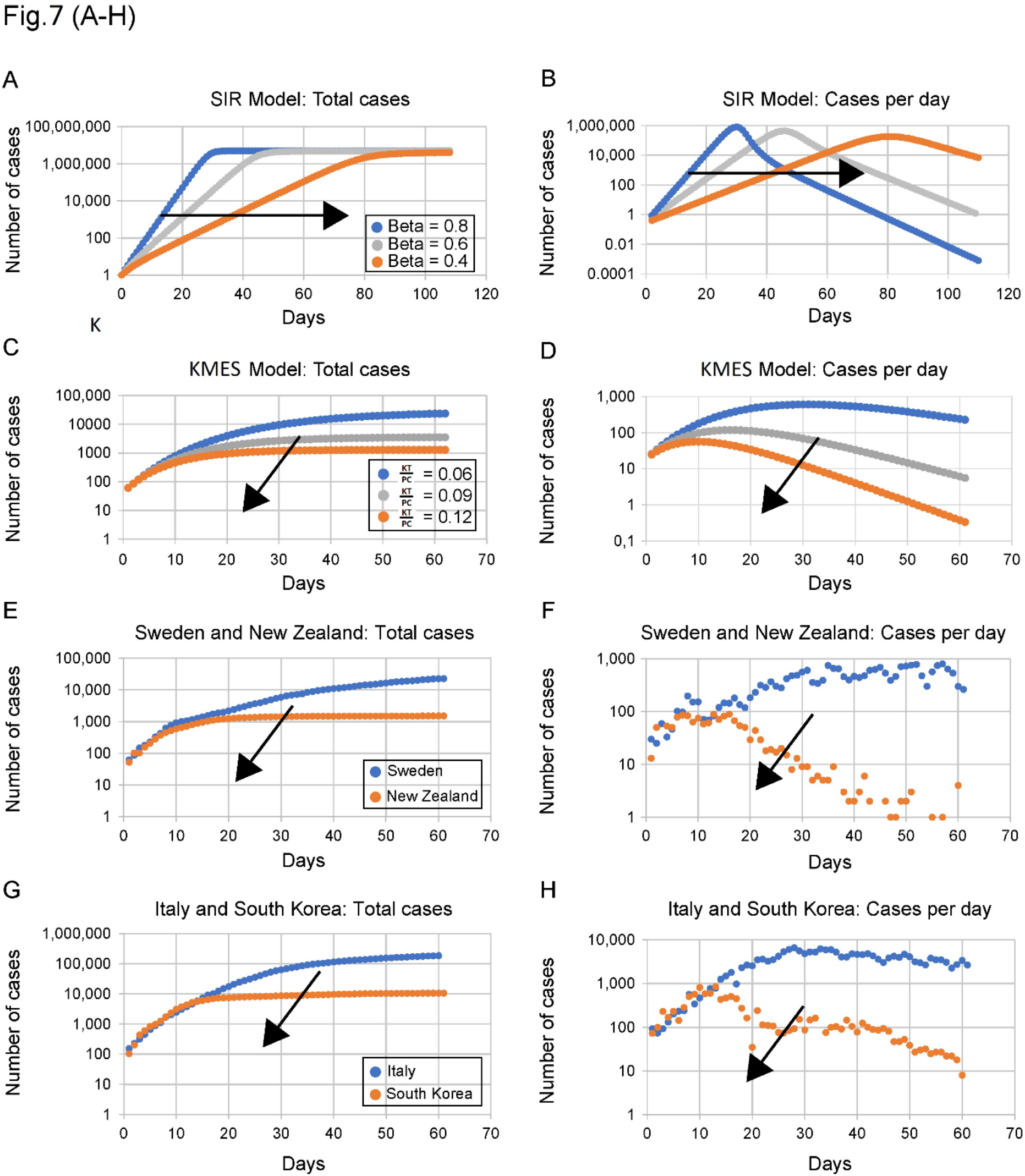
Comparisons of predictions of the approximate SIR and complete KMES models with observed data from four countries. Note: Containment measures increase in all panels from blue to grey to orange dot curves. The arrow on each graph indicates the direction of more social distancing. SIR model trend predictions: (A) Total cases; **(B)** Daily new cases. Rate of contact and transmission (β) decreases with increasing social distancing (from blue to grey to orange curves). Rate of recoveries (γ) = 0.2 for both sets of plots. As β decreases, the daily total of cases increases more slowly and plateaus later (**A**). Daily new infections project to later, but only slightly lower peaks (**B**). **KMES model trend projections: (C)** Total cases; **(D)** Daily new cases. As containment measures increase (higher 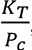blue to grey to orange curves), Equation 23 projects that total cases will rise to lower levels; and reach these levels earlier **(C).** Similarly, Equation 22 projects that new daily cases will peak earlier to lower values with increasing containment **(D).** *K*_*T*_ = 0.2 for plots **(C)** and **(D).** Data reported from different countries during the COVID-19 pandemic. The remaining graphs contain data from pairs of countries with differing containment measures referenced to a day when each member of the pair had nearly equal numbers of new cases. **(E)** Total cases in Sweden (no containment measures, blue) and New Zealand (strict containment, orange).**(F)** Daily new cases in Sweden (blue) and New Zealand (orange).**(G)** Total cases in Italy (loose containment measures, blue) and South Korea (strict containment, orange). **(H)** Daily new cases in Italy (blue) and South Korea (orange). The trends in the observed data, panels (**E – H)**, are the opposite of those exhibited by the ASIR model for increasing containment (decreasing β) in panels (**A, B**). The SIR model trends in (**A**) and (**B**) have completely different shapes; and vary with increasing containment in an opposite sense to those in the country data. The KMES model trends in **(C)** and **(D)** are highly similar to those in the country data **(E – H)**.

We can also mathematically compare the trends projected by the SIR model with trends predicted by the full Kermack and McKendrick equations using the following expression derived from the KMES:

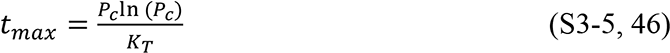

As can be deduced from Equation S3-5, and in contrast to published analytical solutions of the SIR equations (Kroeger and Schlickeiser 2020), the KMES mathematically predicts that the time of the peak in daily cases will occur earlier with increased social containment (i.e., higher 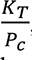). Therefore, SIR projections differ qualitatively from those of the KMES both graphically and mathematically.

The simplest test of the utility of a model is whether it projects the trends actually present in real data. If the projected trends emulate those found in reality, then free parameters within the model are plausibly of value in achieving a higher degree of fit and utility. It is fortuitous, then, that the initial progression of the COVID-19 pandemic was well documented in multiple countries which took different paths while attempting to contain the spread of the virus. This dataset affords the opportunity to test the veracity of the trends predicted by both the SIR and KMES models using real data.

In plots E to H in Figure 7, we can compare the SIR and KMES projected trends to COVID-19 pandemic case data (Roser et al 2021) for total cases and for daily new cases in Sweden and New Zealand (Figure 7E and F), and in South Korea and Italy (Figure 7G and H). The paired countries have comparable population densities but implemented mitigation measures with different intensities (Campbell 2020, Field 2020, Orlowski and Goldsmith 2020, and Sanfelici. 2020). New Zealand and South Korea introduced stronger containment measures much earlier than Italy and Sweden.

In support of the KMES and in contradistinction to the SIR model, the country data in Figures 7E–H show that stronger containment measures are associated with an *earlier* levelling off at a *lower* total number of cases and an *earlier* and *lower* peak in new infections. Harris (2023) also reported this phenomena. In Figure 4 (Harris 2023), the peak of the Omicron wave at the beginning of 2022 in large US counties where the mobility had been highly reduced occurred approximately a week earlier than the corresponding peak in counties where the mobility had not been so significantly reduced. In Harris’s data and in Figure 7 above, trends in both peak position and height demonstrate that SIR models are not merely inaccurate, a tolerable trait in an approximation, but project epidemic data to trend in the *opposite* direction to the reported data; a behavior that no amount of free parameter fitting can correct. Therefore, the SIR model both contradicts the KMES <u>and</u> fails the simplest test of model veracity: the projection of qualitative trends.

### 3.2 Understanding the Implications of the “Flatten the Curve”

As seen in the preceding section, though both *β* and *P*_*c*_ are posited to represent social interaction in their respective models, the trend in the movement of the daily cases peak with decreasing social interaction (decreasing *β*) in the SIR model is opposite to that with decreasing social interaction (decreasing *P*_*c*_) in the KMES. Since the KMES reproduces the observed trends and the SIR model does not, it seems likely that the nature and implications of the SIR assumptions may not be sufficiently understood.

To understand these implications, we start by examining the conventional assumption that *φ*(*t*, *θ*) and *Ψ*(*t*, *θ*) can be constants. Using Equations 38 and 39, and the prior definition that 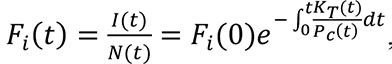, we can find expressions for the time varying *K*_*T*_(*t*) and *P*_*c*_(*t*) when *φ* and *Ψ* are assumed to be the constants *β* and *γ*:

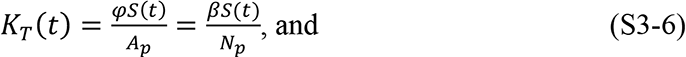

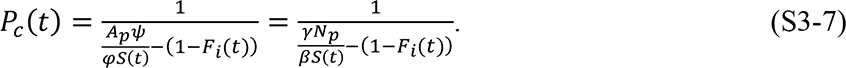

From Equation S3-6 we see that *φ* can only remain constant if *K*_*T*_(*t*) decreases in direct proportion to the decreasing size of the susceptible population, *S*(*t*). This is implausible on its face because, as discussed in the derivation of the solution, *K*_*T*_(*t*) is solely a function of the disease agent and thus, is likely a constant for a substantial time at the beginning of the epidemic; at least until the disease agent itself is modified by mutation or selection. Kermack and McKendrick (1927) themselves, in their introduction on pp. 702, note that it is implausible to assume that disease transmissibility will *decrease* as the disease spreads.

Furthermore, even if transmission were to decrease over time within the infected population, it is improbable that this decrease would, as required by Equation S3-6, occur in a fixed linear proportion to the remaining number of susceptible people. Thus, the assumption within the SIR model that *φ* can be modelled as a constant requires an implausible additional assumption.

It is not possible to state how *P*_*c*_(*t*) must vary to maintain *Ψ* as a constant by merely inspecting Equation S3-7. We can, however, extricate the behavior of *P*_*c*_(*t*) required by Equation S3-7 by plotting the time series of Equation S3-7. To accomplish this, we first simulate the time series of both *N*(*t*) and 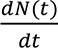 using an Euler approximation of Equations 16 and 43 and a time step of 0.1 day. Using the same values of *β* and *γ* employed in Figure 7A and B, Equations S3-6 and S3-7 were then used to determine the values of *K*_*T*_(*t*) and *P*_*c*_(*t*) at each successive time step.

The purpose of this simulation was first to demonstrate that imposing the conditions of Equations S3-6 and S3-7 on *K*_*T*_(*t*) and *P*_*c*_(*t*) will enable the KMES to produce the same results as the SIR approximation. The second purpose was to determine and demonstrate the actual implausible temporal behavior the SIR approximation imposes on both *K*_*T*_(*t*) and *P*_*c*_(*t*).

The time series plots of the simulation of both *N*(*t*) (cases) and 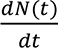 (cases per day) appear in Figure 8. The close approximation of the KMES and the SIR curves in Figure 8 demonstrates that the SIR model is, indeed, a subset of the KMES when the constraints of Equations S3-6 and S3-7 are applied to *K*_*T*_(*t*) and *P*_*c*_(*t*).

**Figure 8.**
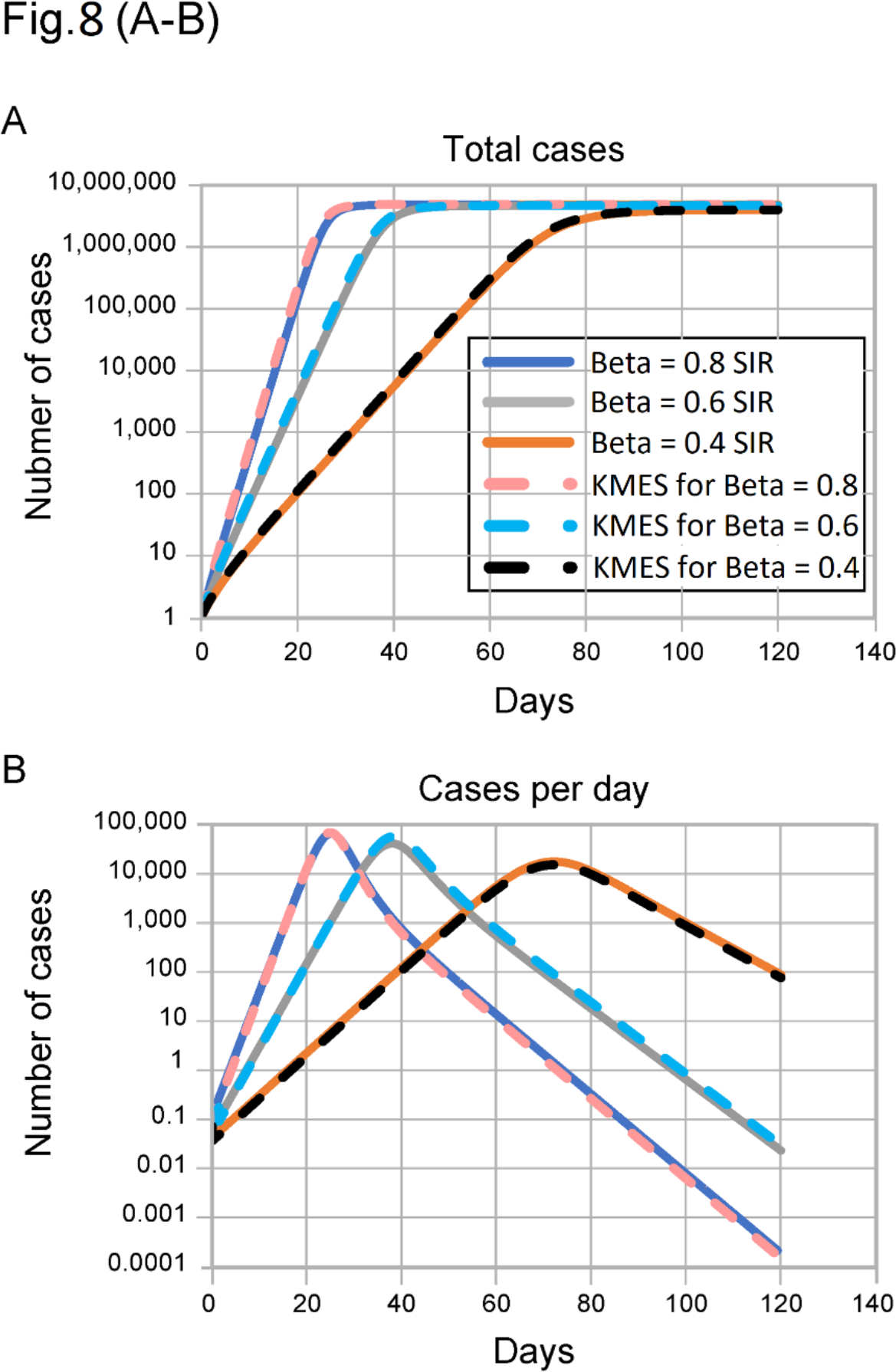
Demonstration that KMES can be modified to produce the SIR model results. Both plots contain 6 lines. For the same β and γ, both the SIR and KMES simulations overlay each other. The KMES simulations were produced by imposing the criteria in Equations S3-6 and S3-7, connecting the SIR and KMES constants. This plot demonstrates that the SIR approximation provides the same result as the KMES provided the constraints of these equations are imposed. Γ is 0.2 for all plots. These plots are the same as the SIR plots in figure7A and 7B on a log scale.

The values of *K*_*T*_(*t*)and *P*_*c*_(*t*) that produce the KMES curves in Figure 8, are plotted in Figure 9. The figure shows that the constraints on *K*_*T*_(*t*)and *P*_*c*_(*t*), imposed by the SIR model, compel the acceptance of unlikely phenomena; namely, that *K*_*T*_(*t*) decreases with time and *P*_*c*_(*t*) must behave in an unrealistic manner.

**Figure 9.**
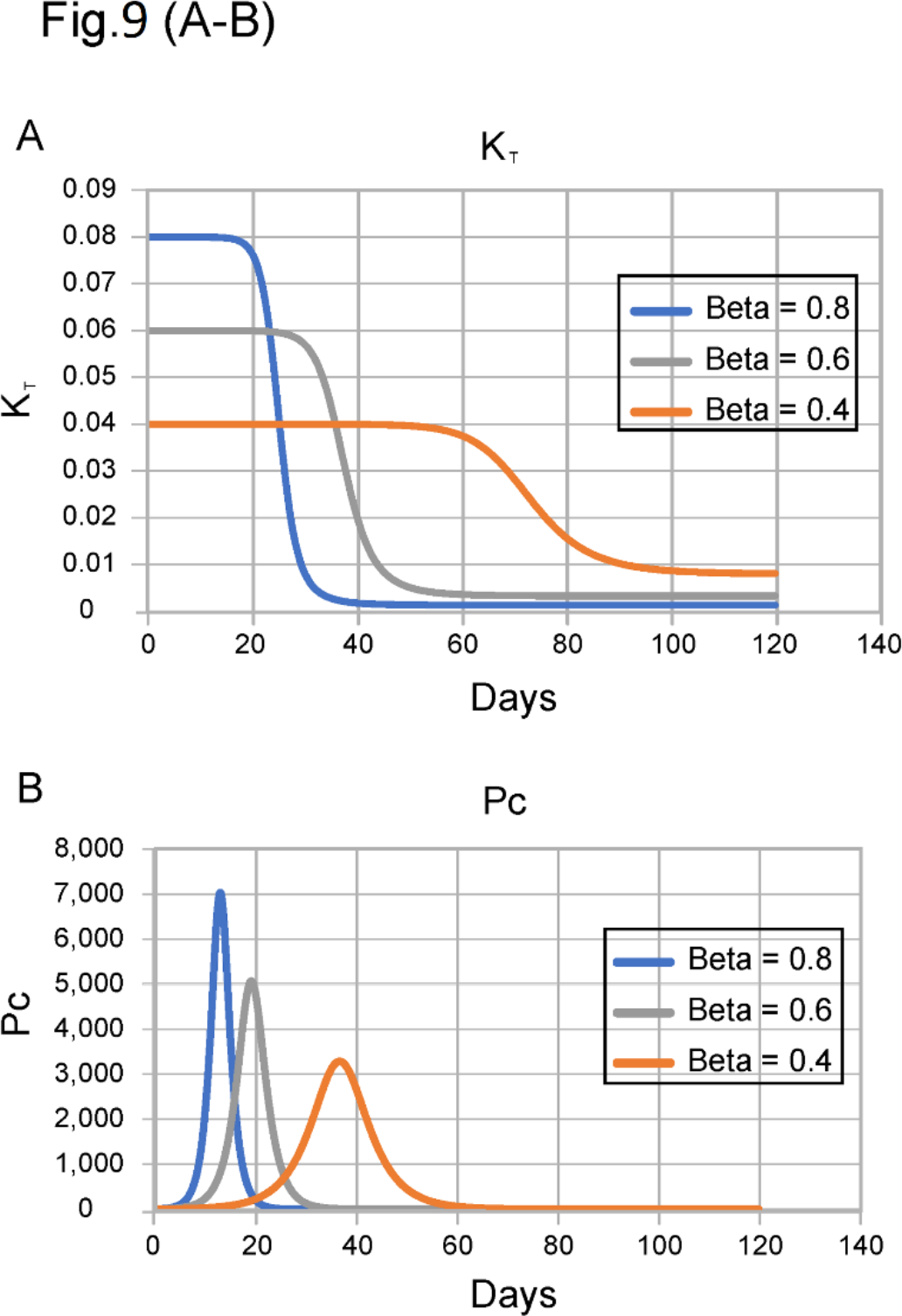
Time series for creating solution curves. These graphs show how Pc and KT are forced to vary within the KMES simulation shown in Figure 8 when the constraints of Equations S3-6 and S3-7 (constant φ and ψ) are imposed. Γ is 0.2 for all plots. X-axis is days.

As Figure 9B illustrates, the consequent, implicit assumption of applying the SIR approximation is that, early in the epidemic, the population increases its contacts, and then suddenly and symmetrically (in time), reverses course and reduces the number of contacts. At each value of *β*, this up and down spike in contacts (Figure 9B) precedes a plunge in the value of *K*_*T*_(*t*) (Figure 9A), and the steep decline is immediately followed by the peak in daily cases (seen in Figure 8B), tailing to the eventual end of the epidemic.

Just as we previewed in the introduction, these implied consequences of a constant *β* in the SIR model make the clear points that a constant *β* does *not* represent constant social interaction; and a higher *β* does *not* represent a consistently higher level of social interaction. Also, a symmetric spike in social interaction (*P*_*c*_(*t*)), higher and earlier, proportional to the value of *β*, followed by an immediate collapse in transmissibility (*K*_*T*_(*t*)), is simply unfathomable.

The consequences of the approximations in the SIR model become even more clear when we make manifest the time varying nature of *β*(*t*) and *γ*(*t*) (and therefore of *φ*(*t*) and *Ψ*(*t*)) required when the quantities *K*_*T*_ and *P*_*c*_ are held constant. Like the preceding analysis, we explored these consequences using simulations of the KMES and the SIR model. Since the KMES predicts the country data well, we also compared the simulation results to two of the country results (Italy and New Zealand).

As a first step, we used the values of *ln*(*F*_*i*_(0)*K*_*T*_) and 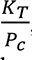 in Table 1 to derive values of 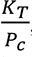 and *K*_*T*_ for Italy and New Zealand and used the solution to project the results. We then simulated the SIR model with the assumption that the values of *β* and *γ* (and therefore *φ* and *Ψ*) were constant and equal to the values of 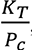 and *K*_*T*_ used in the solution. The results of both the KMES and SIR simulations are plotted in Figure 10, along with the country data. As can be easily seen, the KMES model accurately models the country data, and the SIR model does not.

**Figure 10.**
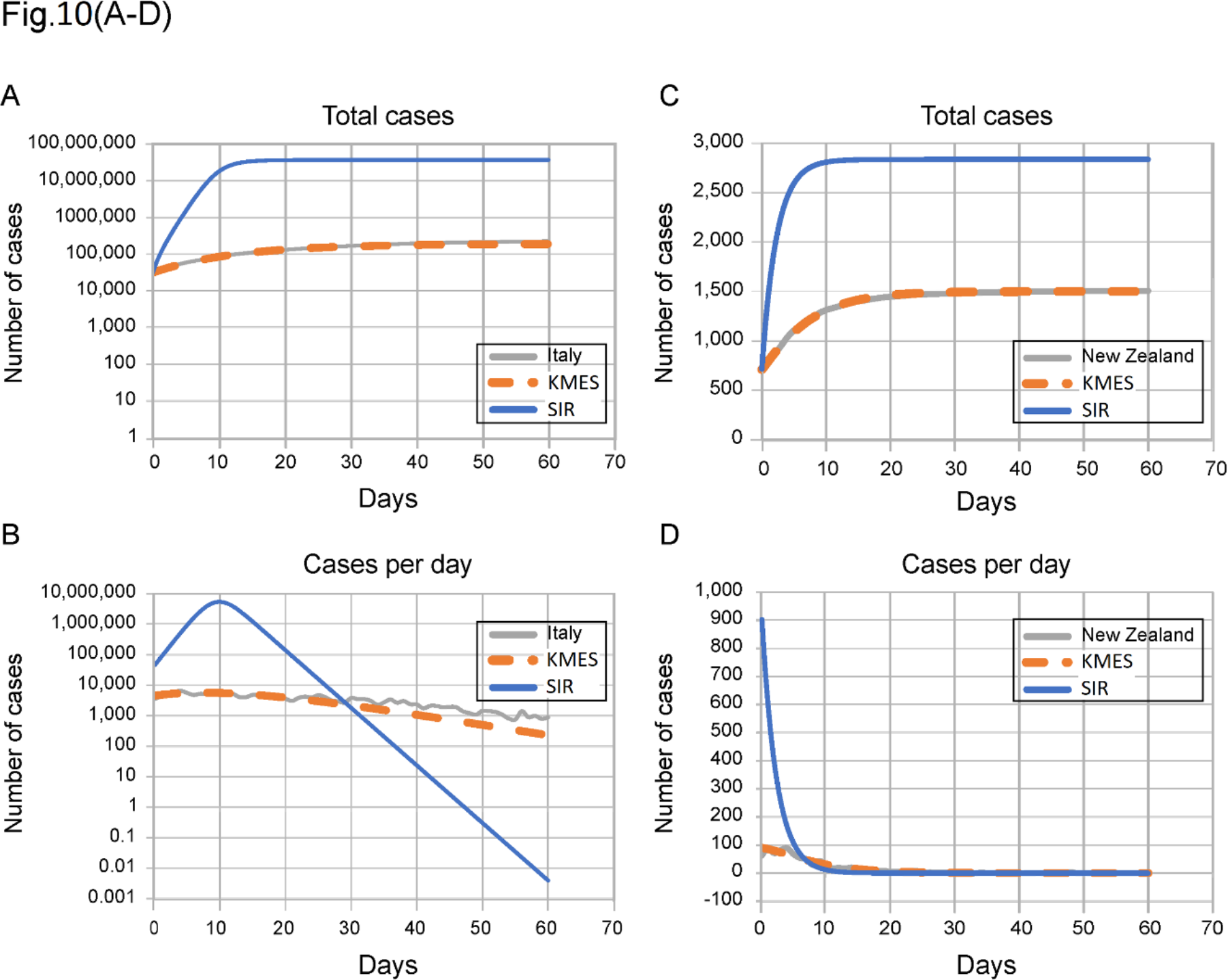
SIR and solution simulations of the Italy (10A & B) and New Zealand (10C & D) data. 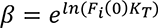 where *ln*(*F*_*i*_(0)*K*_*T*_) is from Table 1 and *γ* is equal to the 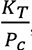 constant from Table 1.

In a second step, we recast Equations S3-6 and S3-7 in terms of *β*(*t*) and *γ*(*t*):

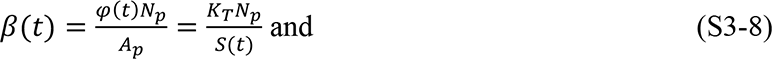

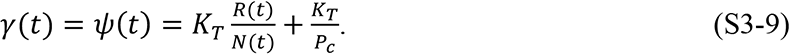

Using Equations S3-8 and S3-9, we then calculated the time series of *β*(*t*) and *γ*(*t*) necessary to generate the KMES curves in Figure 10. Those time series, plotted in Figure 11, show that under the conditions present in the countries, *β*(*t*) is nearly a constant, because, early in the epidemic when *S*(*t*) ≈ *N*_3_, *β*(*t*) ≈ *K*_*T*_, and *β*(*t*) can be approximated as a constant, while *γ*(*t*) clearly is not constant.

**Figure 11.**
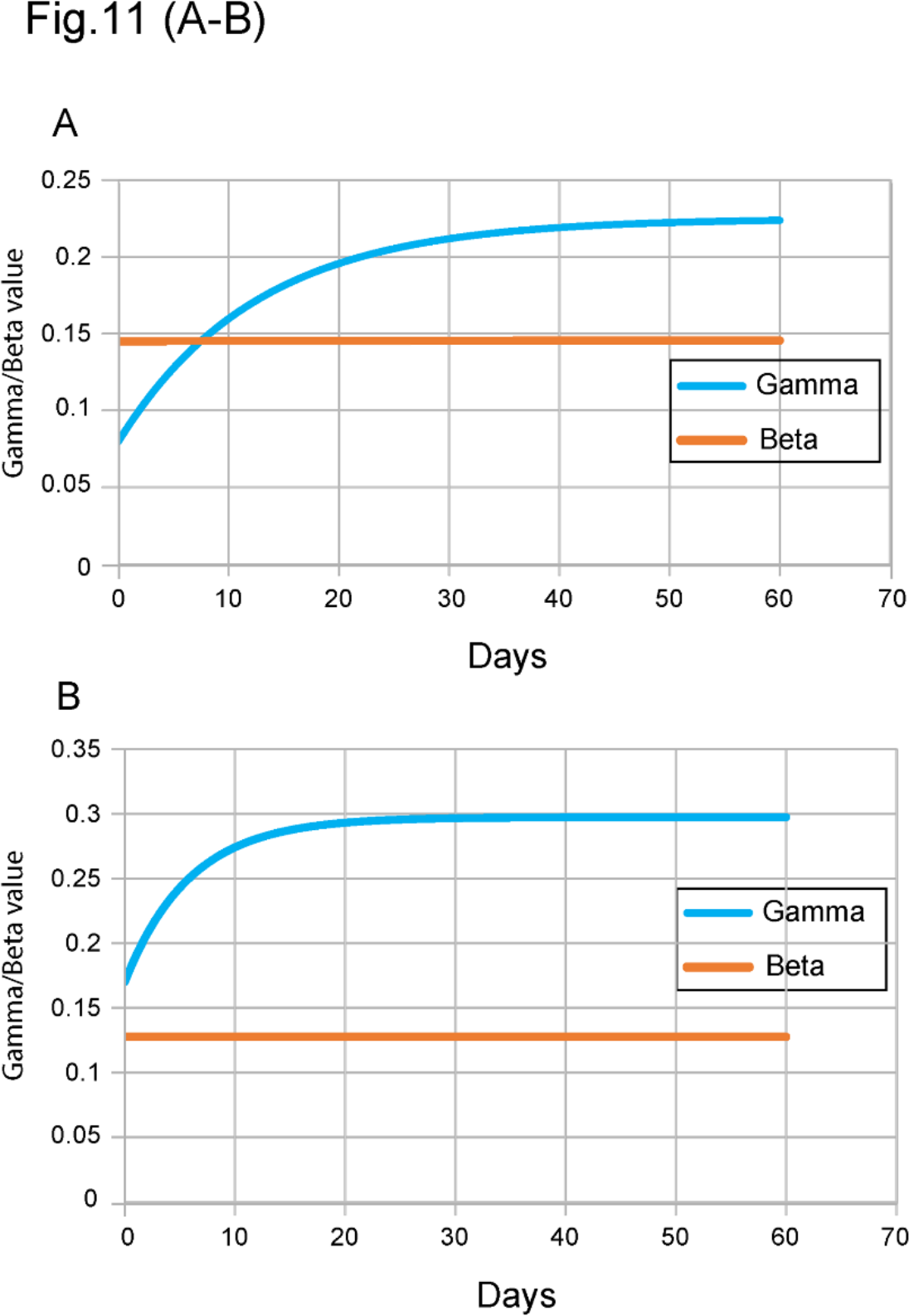
Time series for *γ* and *β*. A) Italy. **B)** New Zealand. These are the values of *γ* and *β* necessary for the SIR approximation to accurately model the country data.

As a last step in the analysis, we used the values of *β*(*t*) and *γ*(*t*) plotted in Figure 11 in the SIR model to generate the curves in Figure 12. This figure shows that when *β* and *γ* are forced to vary according to Equations S3-8 and S3-9, the SIR model fits the country data quite well.

**Figure 12.**
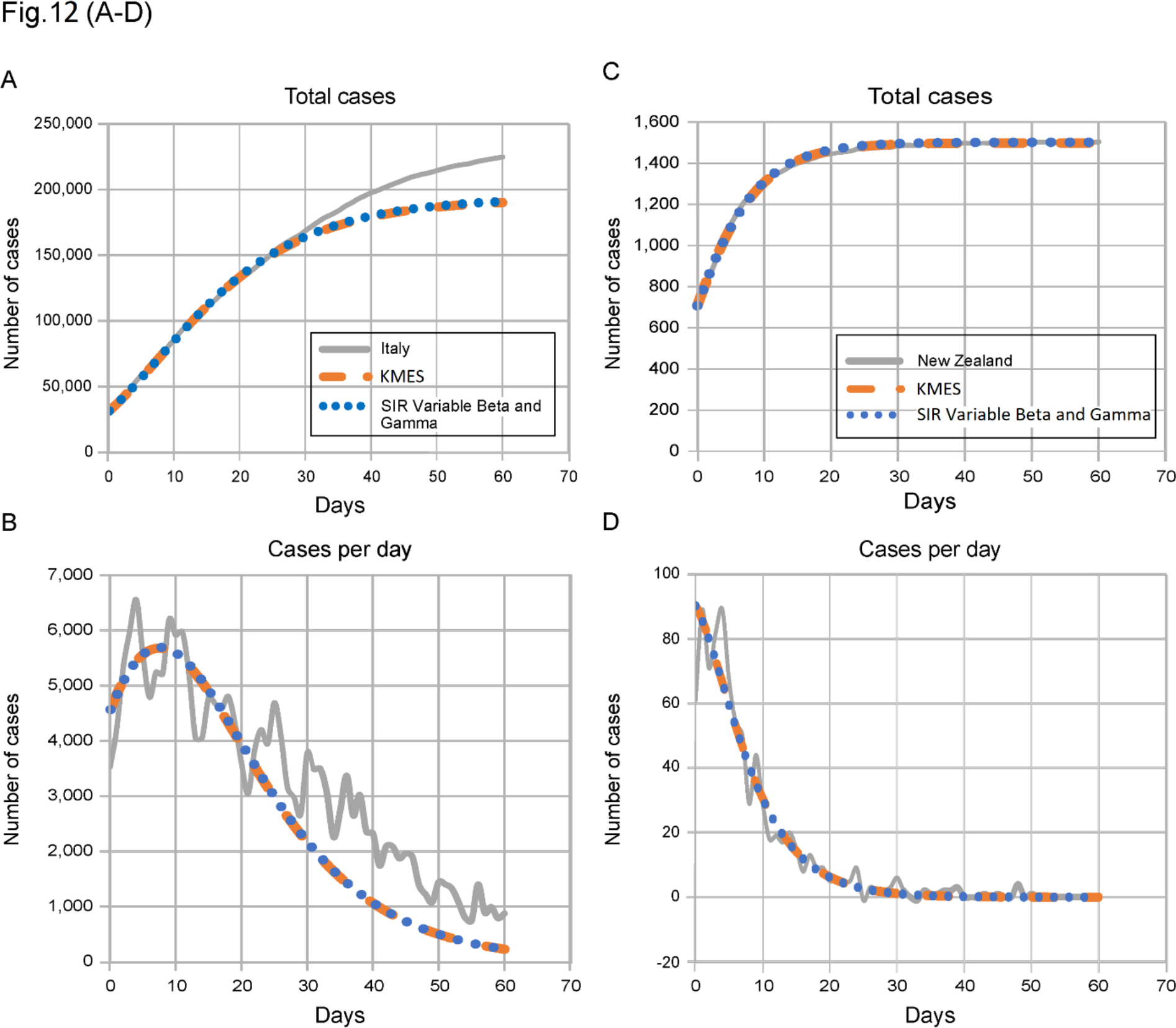
Total cases and new cases per day for Italy and New Zealand. The country data and KMES plots are the same as in Figure 10. The SIR Variable *β* and *γ* plot uses the *β* and *γ* values from Figure 10 in the SIR equations.

Figures 8 and 12 provide another validation of the veracity of the solution. In Figure 8, we show that the solution can be configured to replicate SIR simulations by embedding the SIR approximations within the solution framework. In that setting, the two piecewise and logically invariant solution parameters, KT and Pc, are forced to take implausible and unrealistic time courses. Figure 12 demonstrates, in counterpoint, that an SIR model can produce results identical to the KMES if *β* and γ, the approximations of *φ*(*t*, *θ*) and *Ψ*(*t*, *θ*), are permitted to vary in time according to Equations S3-8 and S3-9. The SIR model fits reality only when “coached” to a time variation for its two parameters using insight derived from the KMES. The foregoing discussion has utilized one of the simplest models of the SIR types; nevertheless, the conclusions apply to all the variations of SIR models.

## Supplement 4. Controlling epidemics early

The quantitative mathematical relationships derived from the KMES in Section 1 characterize the dynamics of an epidemic and illustrate that strong and early intervention is critical. Equation 45 quantifies that the ultimate number of individuals infected in an epidemic, *N∞*, will be exponentially dependent on the number of people with whom each person interacts.

The real-world country data provide vivid examples of the consequences projected by the KMES. Both South Korea and New Zealand enacted strong and early interventions compared to other countries (5,6), as reflected by their 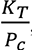 values (Table 1). These strong interventions led to earlier peaks in new cases and to far fewer total cases than in other countries (Figures 2 and 3) in the first few months of the pandemic: the peak number of new cases in both South Korea and New Zealand was 90–99% lower than in other countries, a compelling validation of the explicit statement in the KMES that strong intervention leads to *exponentially* more favorable outcomes.

In the USA, interventions initiated on March 16 began to have an effect around March 23, 2020 (Figure 3B); the number of active cases on March 23, 2020 (Roser et al 2021) was 46,136 (Table 1). Using the values of ln (*F*_*i*_(0)*K*_*T*_(t)) and 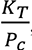 from Table 1, Equation 45 predicts that the ultimate number of cases would have been approximately 1.22 million. If the same intervention had been implemented and sustained starting on March 10, when there were 59 times fewer (782) cases (Roser et al 2021), the model predicts that the ultimate number of cases would also have been 59 times lower, or 20,725. Thus, earlier action could have reduced the ultimate number of projected cases by more than 98%. Of course, the projected estimate of approximately 1.22 million total USA cases would only have occurred if the effectiveness of the interventions that were launched on March 16 had been sustained. Unfortunately, a marked reduction in effective interventions occurred in many parts of the USA in mid-April, well before the official reopening of the economy (Elasser 2020). This caused a second surge in new cases in late April and is why the observed data and the model prediction diverge in Figure 3B.

As shown in the main body of the paper, Section 2, the KMES provides an estimate of the time to the peak of new cases, *t*max. Using Equation 50 and the values of ln (*F*_*i*_(0)*K*_*T*_(t)) and 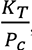from Table 1, the predicted peak in new cases in the USA would have occurred near March 24 if the intervention had begun on March 10. Instead, a 6-day delay in effective intervention shifted the initial peak to April 11, 16 days later, as projected, and that peak was much higher (Figure 3B).

As shown, too, in Section 2, epidemic acceleration, the instantaneous potential to change the pace of the epidemic, can be determined at any point in the epidemic and depends on the social containment actions in effect at that time (Equation 53). What is perhaps less apparent, but predicted by the KMES, is that two countries with identical numbers of cases on a given day can, in fact, have different accelerations on the same day, and will, therefore, exhibit different dynamics immediately after that day.

South Korea and New Zealand (Figure 2A and F) had nearly identical case counts when each imposed strong containment measures (204 cases in South Korea on February 21, and 205 in New Zealand on March 25). Their models suggest that their interventions were about equally effective (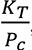 in South Korea and 0.17 in New Zealand; see Table 1). However, since South Korea has a much higher population density than New Zealand ((Worldometers 2021), data in Table 2), it had a much higher number of interactions when the interventions were imposed and, therefore, a higher rate of acceleration, as evidenced by its higher RCO at the time of intervention. Indeed, the rate of change of new cases *was* higher in South Korea than in New Zealand, and the later number of cases in South Korea *was* higher than in New Zealand (Figure 2A and 2F).

Equation 44 clearly illustrates these lessons. As social distancing is strengthened (lower *P*_*c*_), the Effective Replication Number decreases, and the epidemic slows. Early and strong interventions, especially in countries with indigenously high levels of social interaction, are necessary to stop an epidemic in the initial stages. Reopening, enacted too early, can reignite the epidemic, dramatically increasing the number of cases. The astonishing magnitude of the effects, driven by only a few days of delay, derives from the doubly exponential nature of the underlying relationships.

## Supplement 5. Ending an Ongoing Epidemic

We can use the KMES to design measures to end an epidemic in an advanced stage. The management plan is built by first using Equation 57 to predict the number of days a given level of intervention,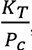 is needed to reduce the new daily cases by a target fraction, *D_tf_*.

For example, using Equation 57, we see that a country targeting a 90% reduction of new cases per day (e.g., from 50,000 to 5,000 cases per day, *D_tf_* = 0.1), can attain its target in about 12 days by imposing a containment level of 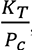 = 0.2. The South Korea and New Zealand data demonstrate that Equation 51 is valid and that 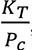 = 0.2 is achievable for this duration. Both countries achieved a value of 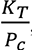 close to 0.2 for the time necessary to produce a 90% reduction.

It took 13 days for South Korea (March 3–16) and 15 days for New Zealand (April 2–15), New Zealand (6) to reduce their new cases by 90% between the dates shown.

Returning to the planning example, after achieving the initial 90% reduction, a reasonable next step might be to relax social containment to a level that allows the economy to remain viable, while preventing the epidemic from erupting again. We can again find the level of 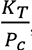 necessary to achieve a chosen target, using Equation 57. If an additional 90% reduction in new cases per day is desired, and a period of 90 days is tolerable for that reduction, then a new level of approximately 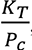 = 0.026 is needed. This equates to a 90-day period during which each person can be in contact with seven specific people, in an infectable way. Note that this is three times *less* stringent than the original USA shutdown level in April 2020 as shown by the level of 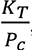 calculated for the United States in that period (Table1). Thus, with a well-planned approach, a country can reduce its new daily cases by 99% in approximately 100 days, enabling the country to control, and essentially end the epidemic, while simultaneously maintaining economic viability.

If even 0.025 is too restrictive, we can choose a still lower 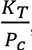 but it must be large enough to avoid a new outbreak. A lower bound for the new value of 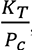 high enough to prevent an outbreak, can be found using Equation 54.

We can easily monitor the progress of interventions using the RCO, as the curve for South Korea illustrates (Figure 1A). Had this country maintained the implemented level of distancing measures, the data would have followed the initial slope. However, the actual data departed from the slope, heralding failures in (or relaxation of) social distancing, which were later documented to have occurred during the indicated time frame (Campbell 2020) (circled data, Figure 1A). Because it summarizes epidemic dynamics, we can use the RCO to continuously determine the effectiveness of implemented measures and whether they need adjustment.

### Supplement 5.1 Outbreaks

We can see from Equation 52 that if the social interventions are strengthened (lower *P*_*c*_) the slope of the RCO curve will steepen and if the interventions are relaxed, the slope will become shallower. Therefore, if the value of *K*_*T*_(*t*) does not change due to a change in the disease transmissibility, the RCO is a metric for monitoring the population interactions. It is also clear that under the assumptions used to develop the KMES, 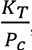 must always be greater than zero, and the RCO slope can never become positive. However, this only remains true if these three conditions remain true: 1) immunity persists, 2) no new infections are introduced from outside the area, 3) Δ*P*_*cni*_(*n*Δ*t*) is a much smaller order of magnitude than the new infections, *K*_*T*_(*n*Δ*t*)Δ*t*. We call the latter two assumptions the assumption that the epidemic is contiguous.

If new infections are introduced into a portion of the population that has thus far been disconnected from the previously infected area, and therefore, has only susceptible people, then the assumption of contiguousness does not hold. This is a common situation when infected people travel from an infected area to a previously uninfected area and cause an outbreak.

In this case, we will begin with Equation 27 and assume that the entirety of the change in *P*_*c*_(*t*) during the time Δ*t* is with uninfected new contacts. That is, Δ*P*_*c*_(*t*) = Δ*P*_*cni*_(*t*); and Equation 27 becomes,

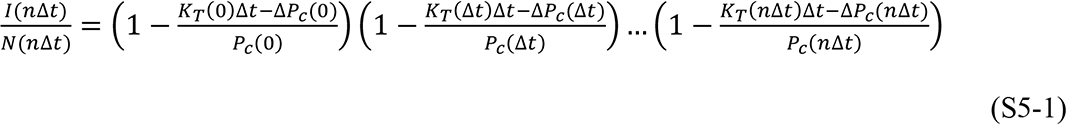

and then, since by definition, *n*Δ*t* = *t*, as *n* → ∞, Δ*t* → 0, Equation 27 becomes,

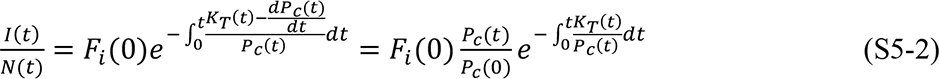

The equations for *N*(*t*), *I*(*t*), and *R*(*t*) are then the following:

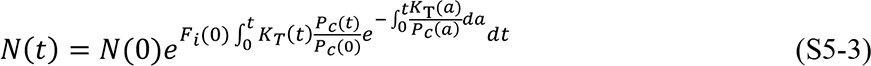

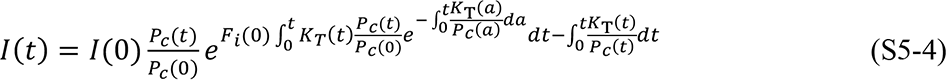

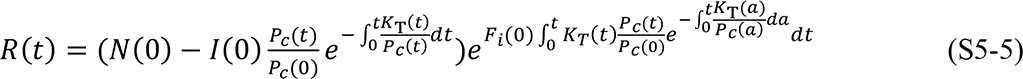

As an alternative to predict the number of cases in an epidemic affected by an outbreak, we can modify Equation 43. Assuming that *t*_-_ = 0, *N*(0) = 1, and introducing the notation *P*_<?_ where *x* denotes the number of the outbreak, Equation 43 can be written as:

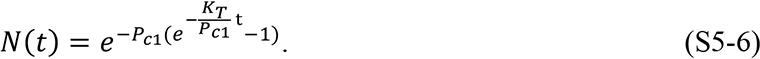

If a new outbreak occurs in a previously unaffected area of a country, then Equation S5-6 can be modified as follows:

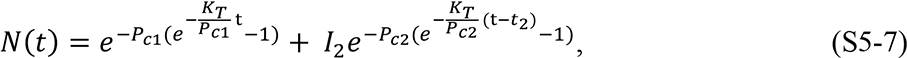

where *N*_Q_is the number of infectious people who initiated the new outbreak, *P*_<Q_ is the social interaction parameter in the new outbreak area, and *t*_Q_ is the time the new outbreak occurs. We have assumed that the disease transmissibility remains the same throughout this illustration. If the transmissibility changes in a subset of the population, then a similar formulation, using the notation, *K*_*T*’?_, can be utilized to track the populations with the new transmissibility.

Equation S5-7 can be written in a general form as

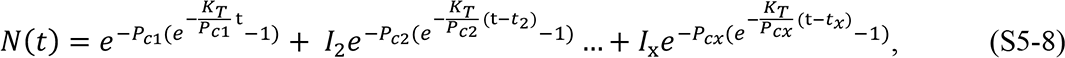

where *x* denotes the outbreak number and t > *t*_Q_ > *t*_l_ > ⋯ > *t*_?_. For each outbreak *t_x_*, *P_cx_*, and *N_x_* need to be determined independently.

While an epidemic is underway, we can detect an outbreak by monitoring the slope of the RCO curve. A positive slope detected in an RCO curve indicates that an outbreak has occurred. This is an indication that immediate action, within days, is required from policy makers to strengthen intervention measures and prevent the outbreak from overwhelming prior progress in controlling the epidemic.

By monitoring the RCO curve, we can also detect if the disease changes its transmissibility through mutation. In this situation, a proper fit of the parameters in Equation 40 is not possible and a modification of *K*_*T*_ is required to accommodate the change.

## Supplement 6: Understanding the Kermack and McKendrick Arrays

The Kermack and McKendrick model in discrete form can be visually represented as an array where the arrays of the variables *N*(*t*, *θ*), *I*(*t*, *θ*), *R*(*t*, *θ*) and their derivatives are defined over t and *θ* according to this array:

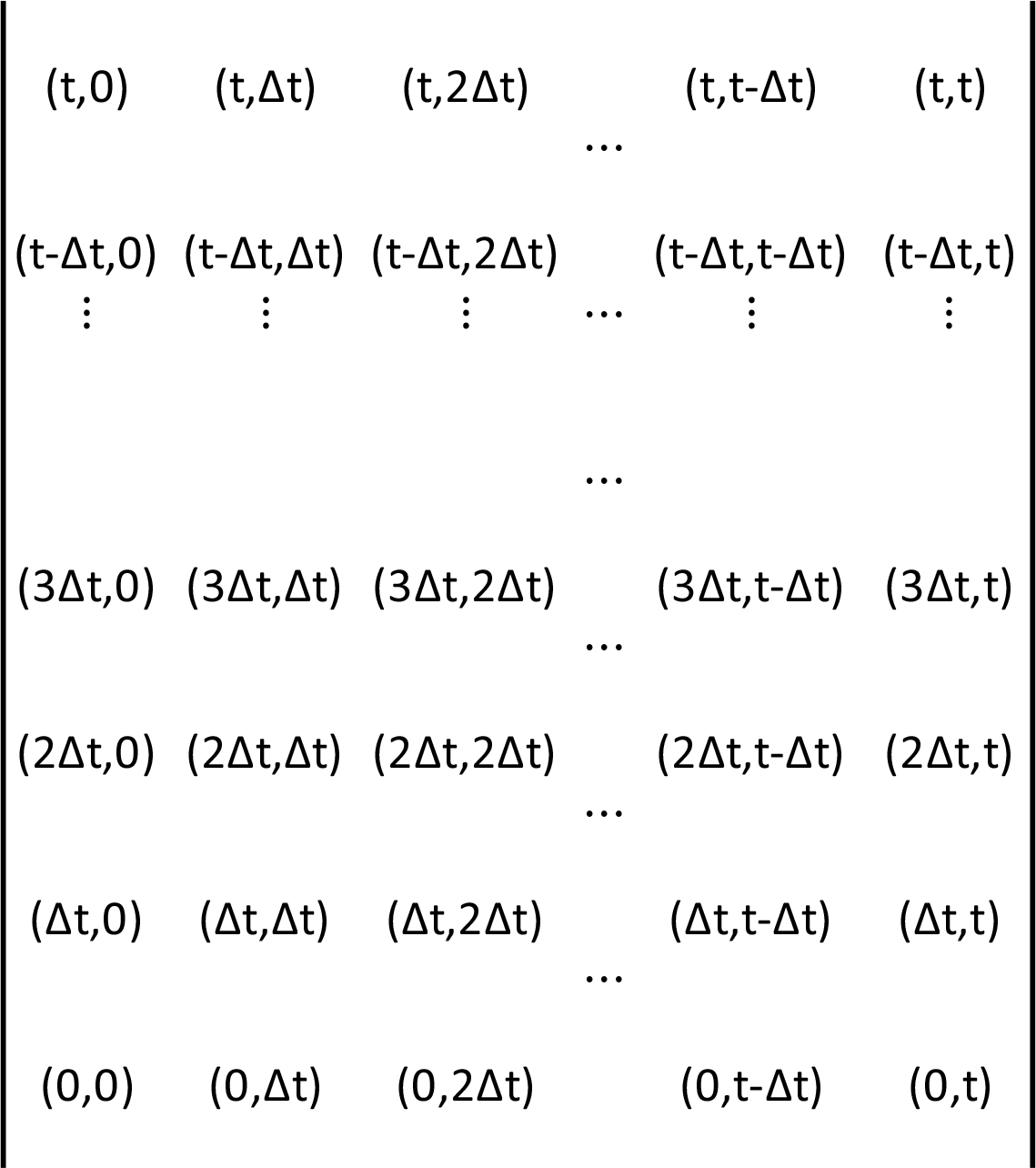

Kermack and McKendrick’s concept of *θ* imagined that the time history of the epidemic was a square t by t array with each row representing an increment of time, Δ*t*, and each column representing the progress of a *θ* group through time. In this conceptualization, each *θ* group starts at time *t* − *θ* and progresses diagonally upward and to the right through the array. This formulation of the problem also means that *θ* has the units of time, *dθ* = *dt*., and Δ*t* = Δ*θ*.

Therefore, Δ*t* and Δ*θ* are used interchangeably throughout the array.

Keeping this convention, we can use Equation 16 and the knowledge that Δ*N*(*t*, *θ*) = 0 when *θ* > 0 to write the matrix for Δ*N*(*t*, *θ*),

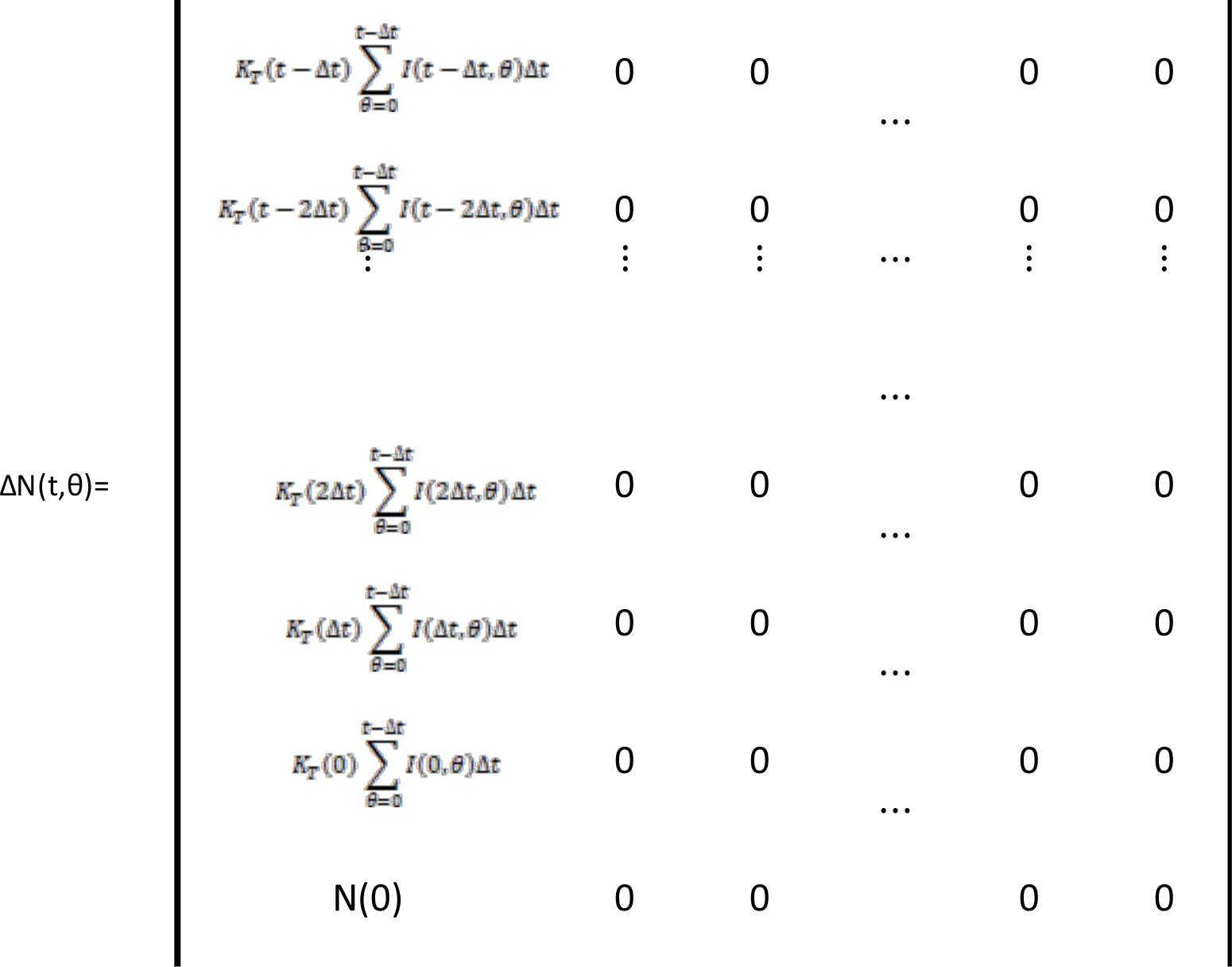

The arrays for Δ*R*(*t*, *θ*), Δ*I*(*t*, *θ*), and *I*(*t*, *θ*) can be written in an identical fashion,

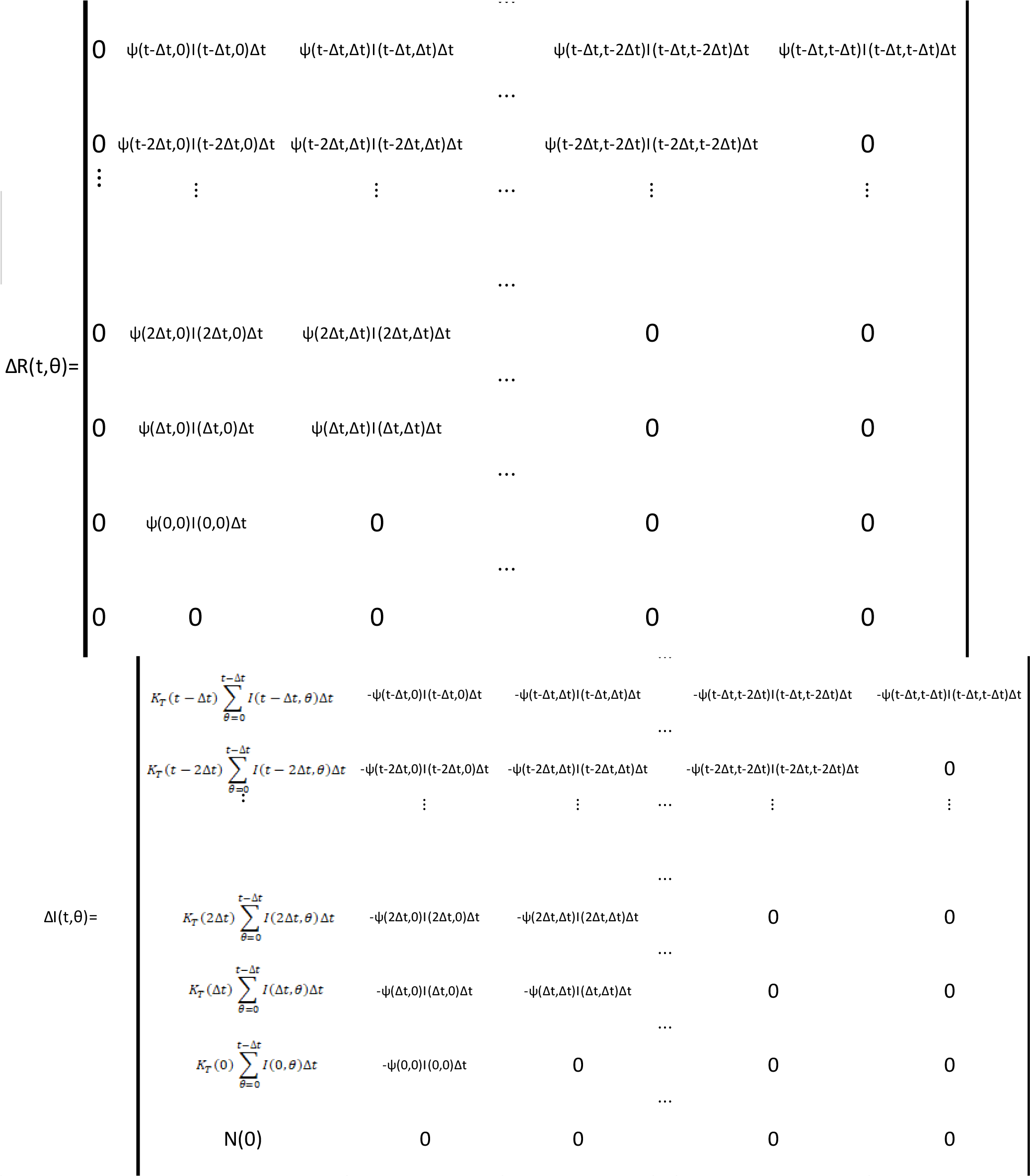

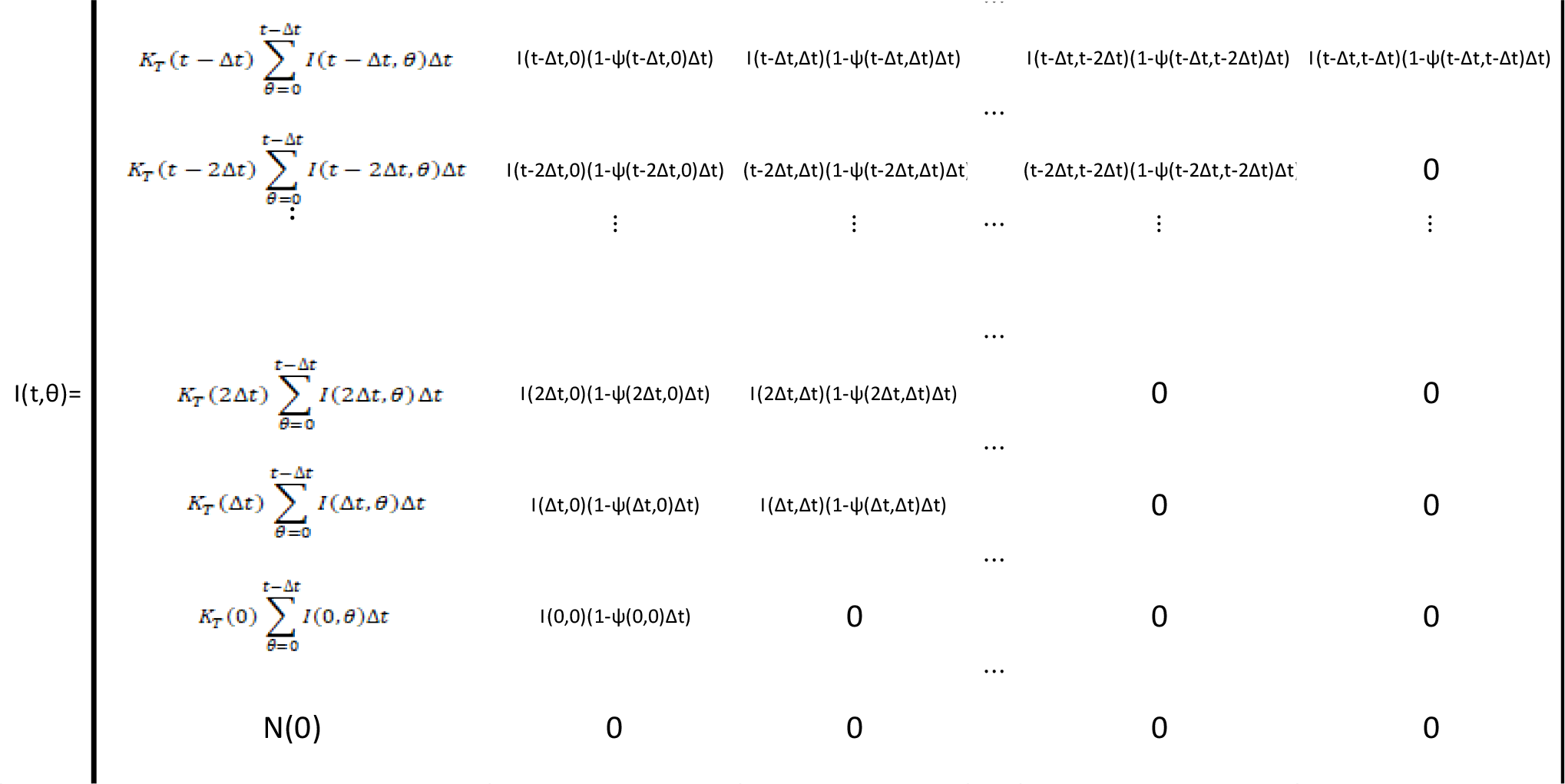

It is clear from this notation that Δ*I*(*t*, *θ*) = Δ*N*(*t*, *θ*) − Δ*R*(*t*, *θ*); and it is interesting to note that 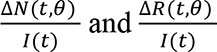 are, respectively, impulse and step functions in *θ*.

We now use the preceding arrays to rederive Equation 3. We with Equation 11,

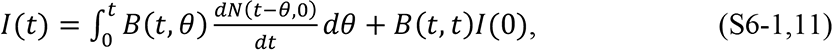

Our goal is to transform Equation S6-1 to Equation 3 to demonstrate that the arrays are a formulation of the Kermack and McKendrick equations.

Referring to the *I*(*t*, *θ*) array and keeping in mind that the value of *I*(*t*, 0) is the value in the (*t*, 0) place in both the Δ*N*(*t*, *θ*) and *I*(*t*, *θ*) arrays, *I*(0, 0) = *N*(0) = *I*(0) as shown in the array, 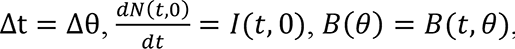, and *B*(*θ*, *θ*) = *B*(*t*, *t*), we know that,

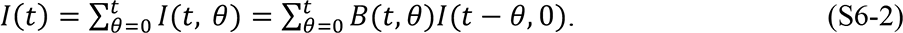

Equation S6-2 is merely the summation form of S6-1. We use the reference to the *I*(*t*, *θ*) matrix to show where it comes from in the (*t*, *θ*) matrices.

Keeping in mind that *Ψ* is only a function of t, it is also clear from the *I*(*t*, *θ*) matrix that,

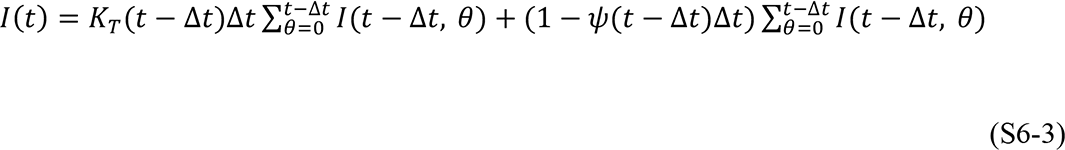

This operation can be repeated all the way back through the matrix to finally obtain the expression,

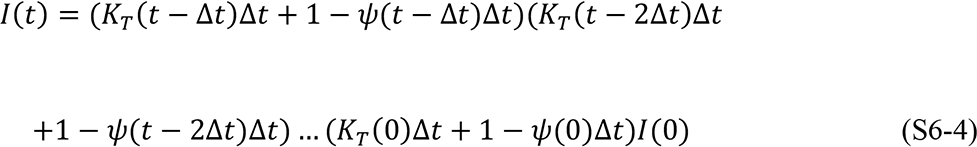

Equating S6-4 with S6-1 and taking the limit as Δt = Δθ go to zero, we arrive at the expression,

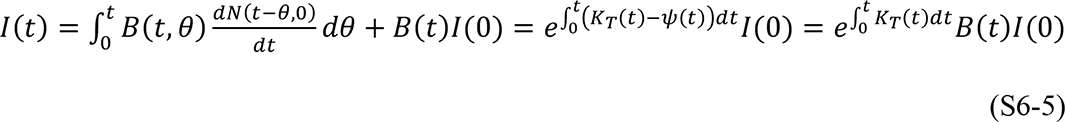

which is identical to Equation 3 with *Ψ*(*t*) = *μ*(*t*). The same approach can be used on the arrays for Δ*N*(*t*, *θ*) and Δ*R*(*t*, *θ*) to find expressions for 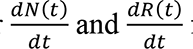 from which the rest of the solution can be determined.

## Supplement 7: List of Equations

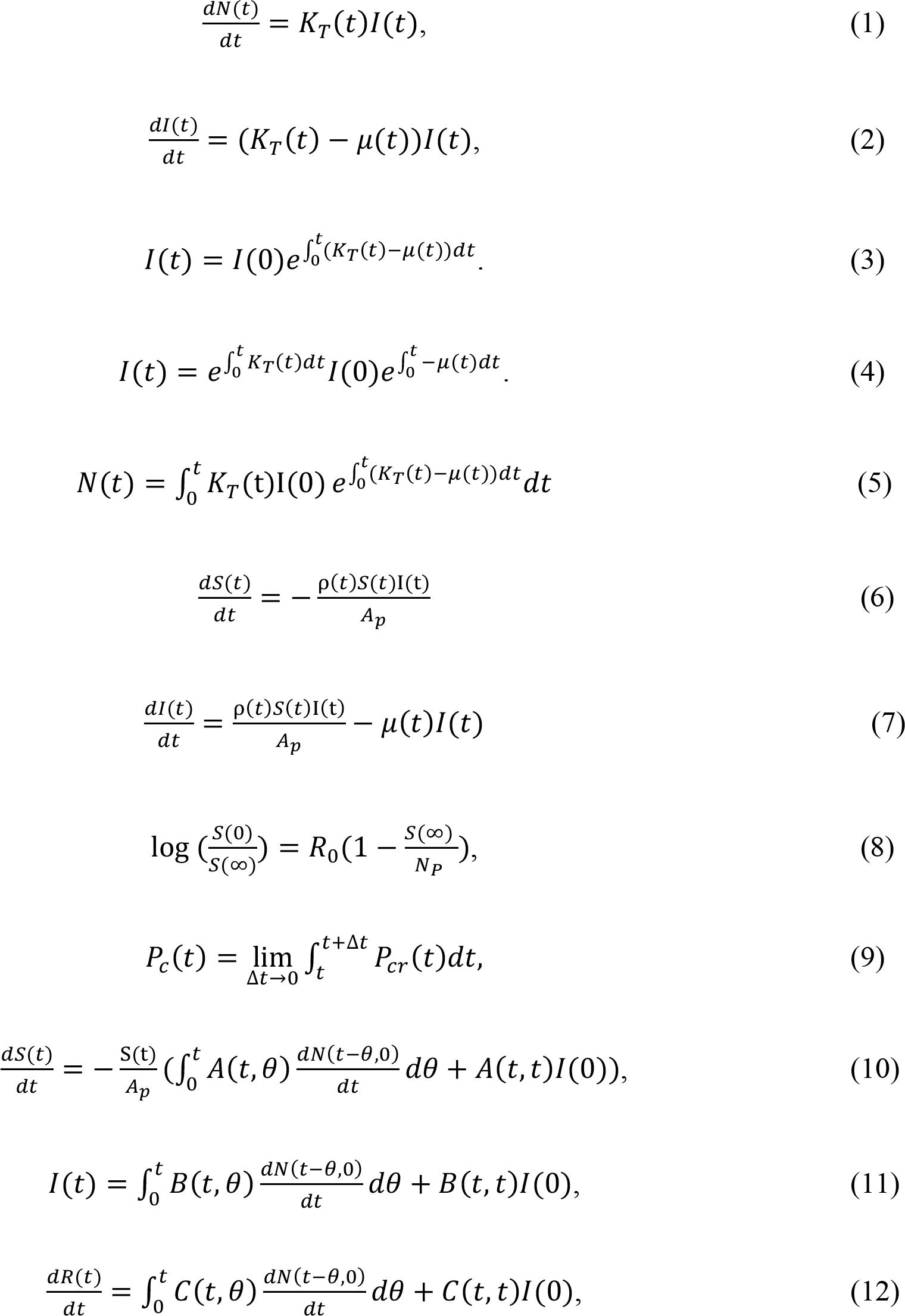
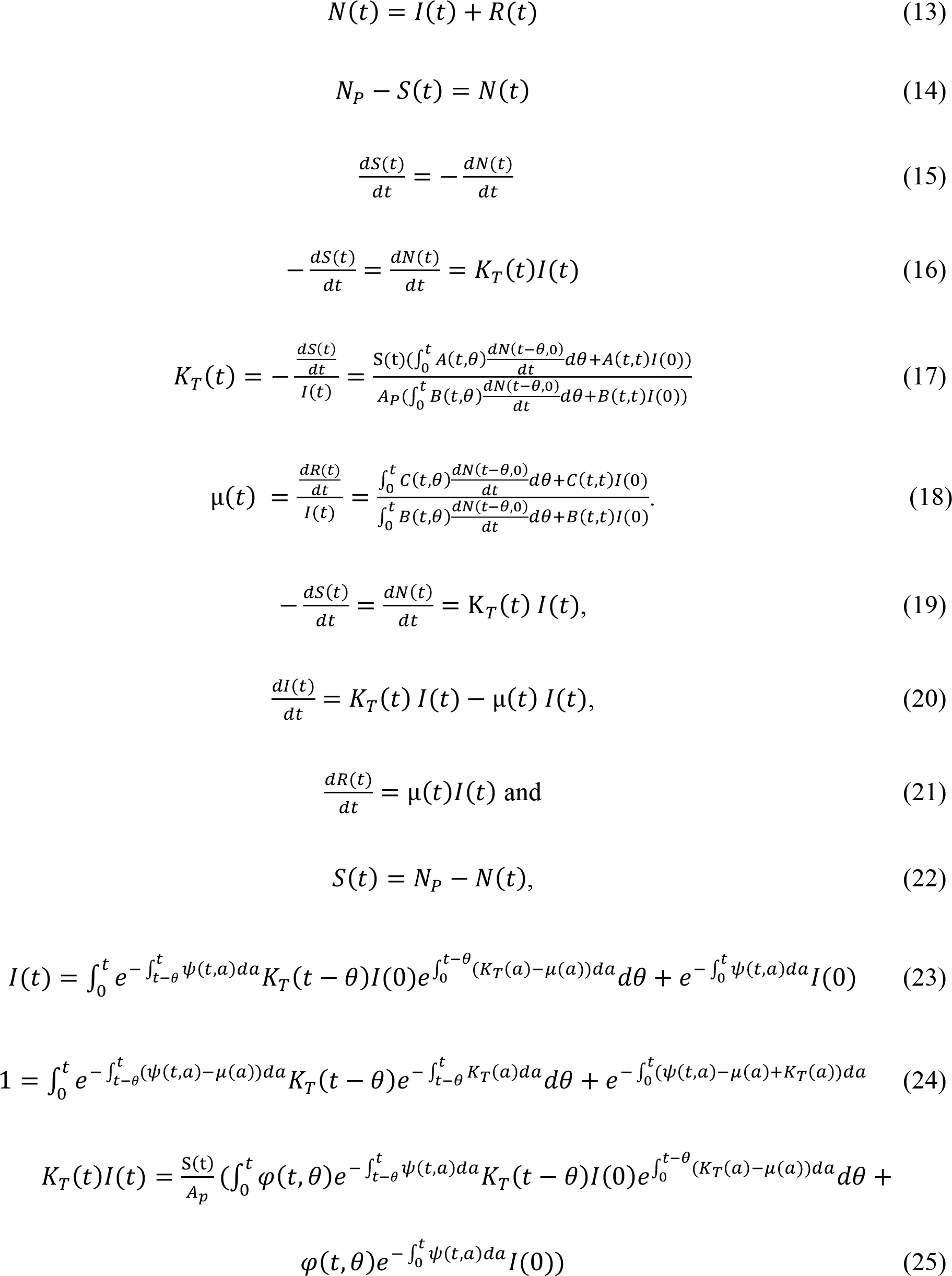
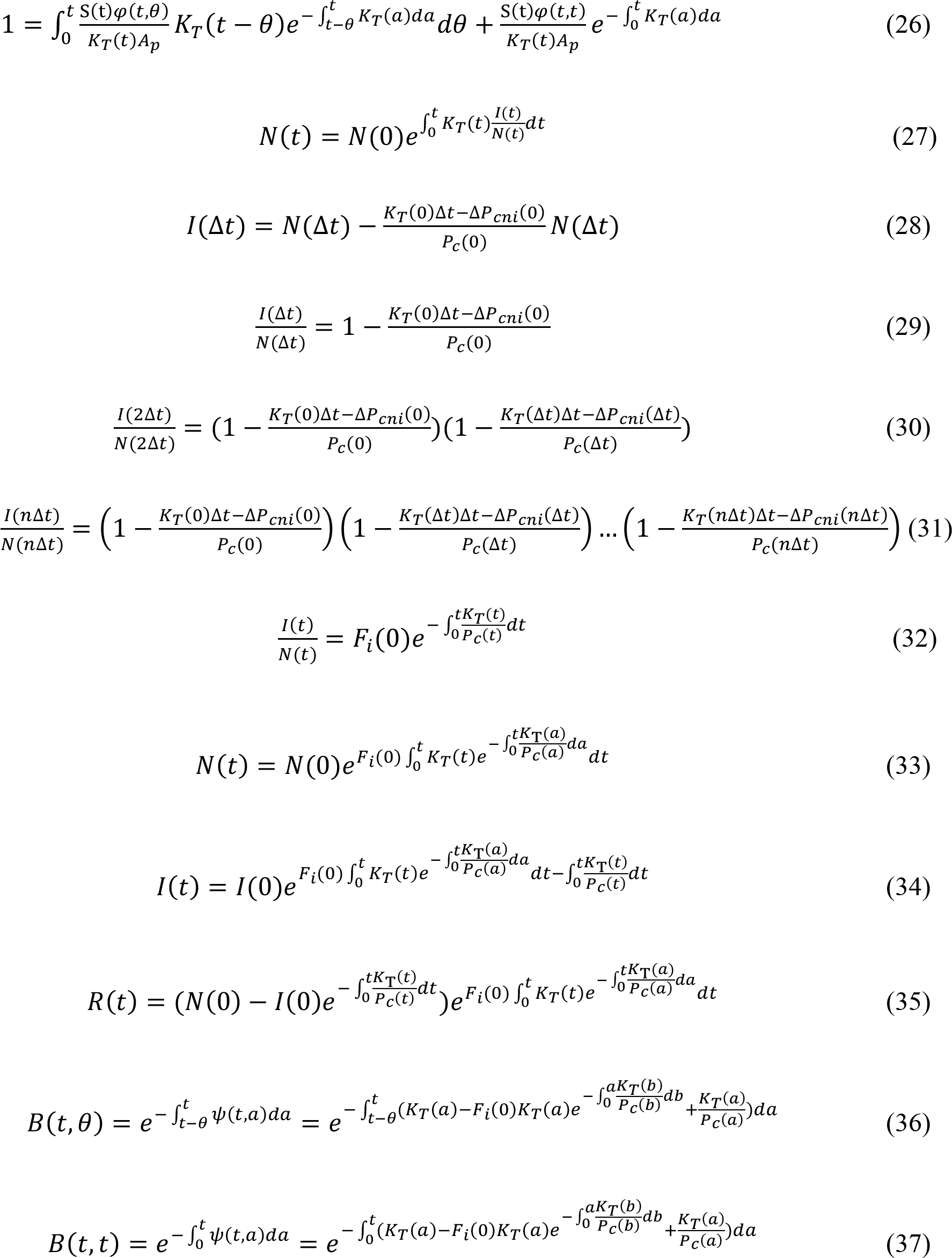
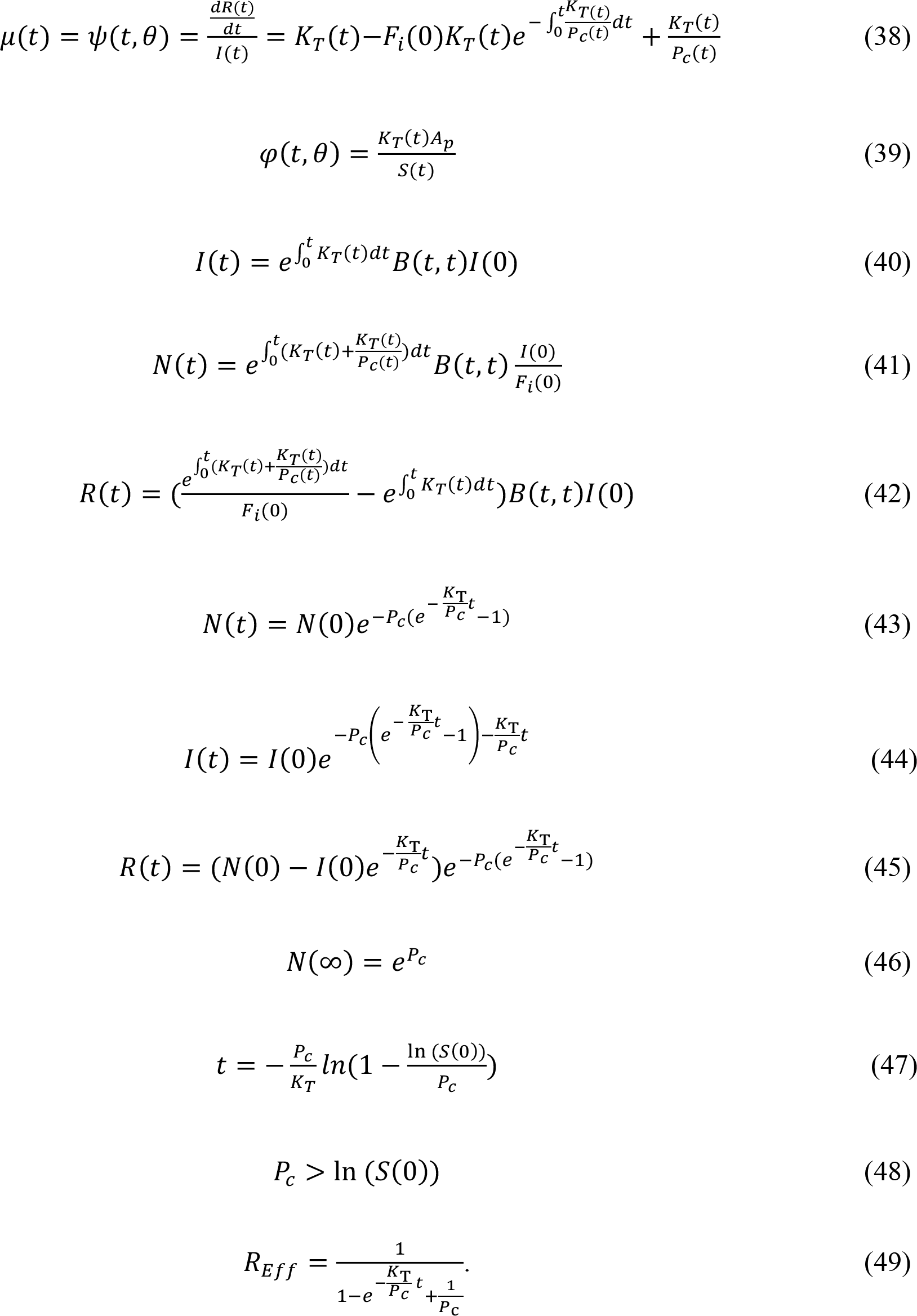
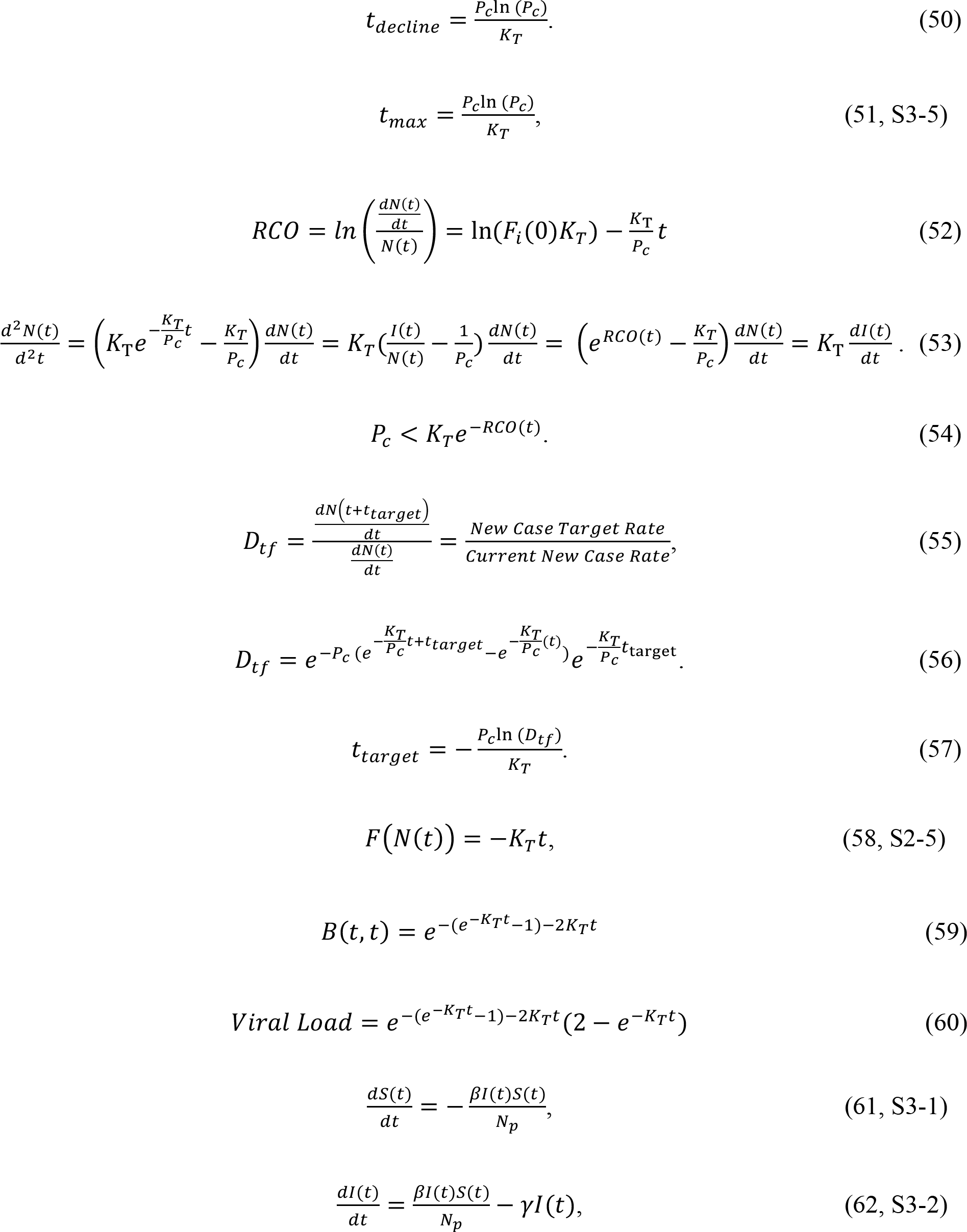
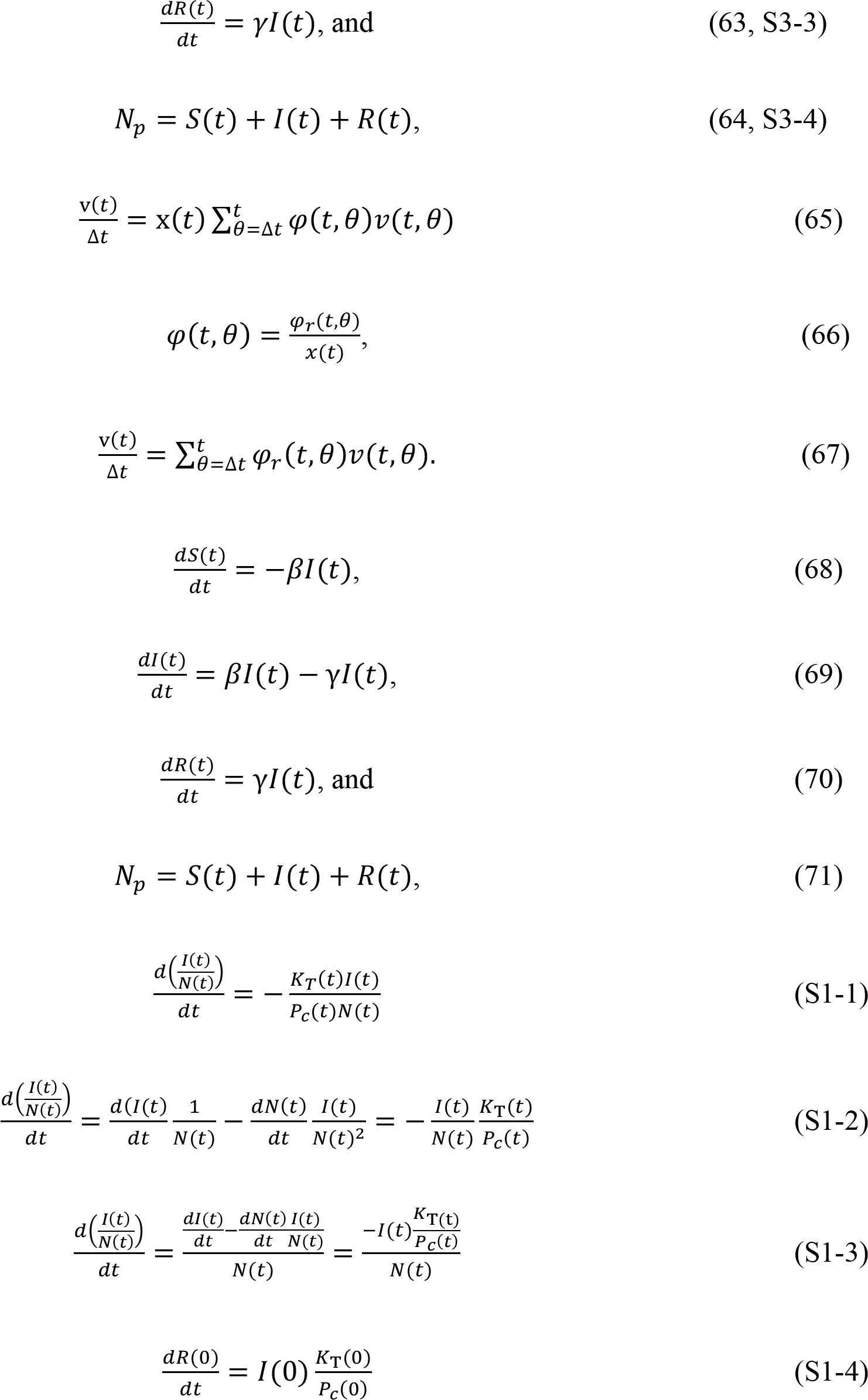
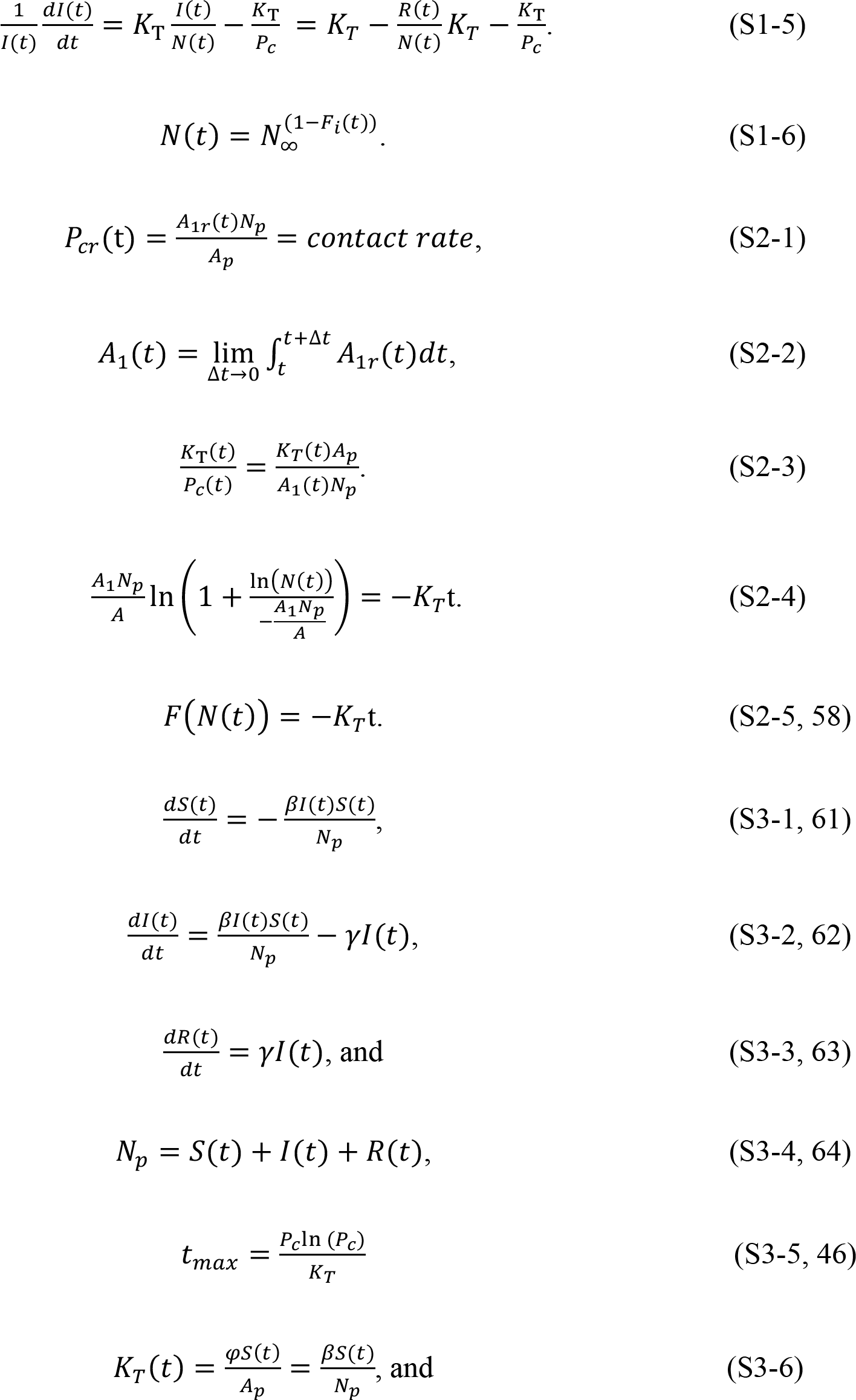
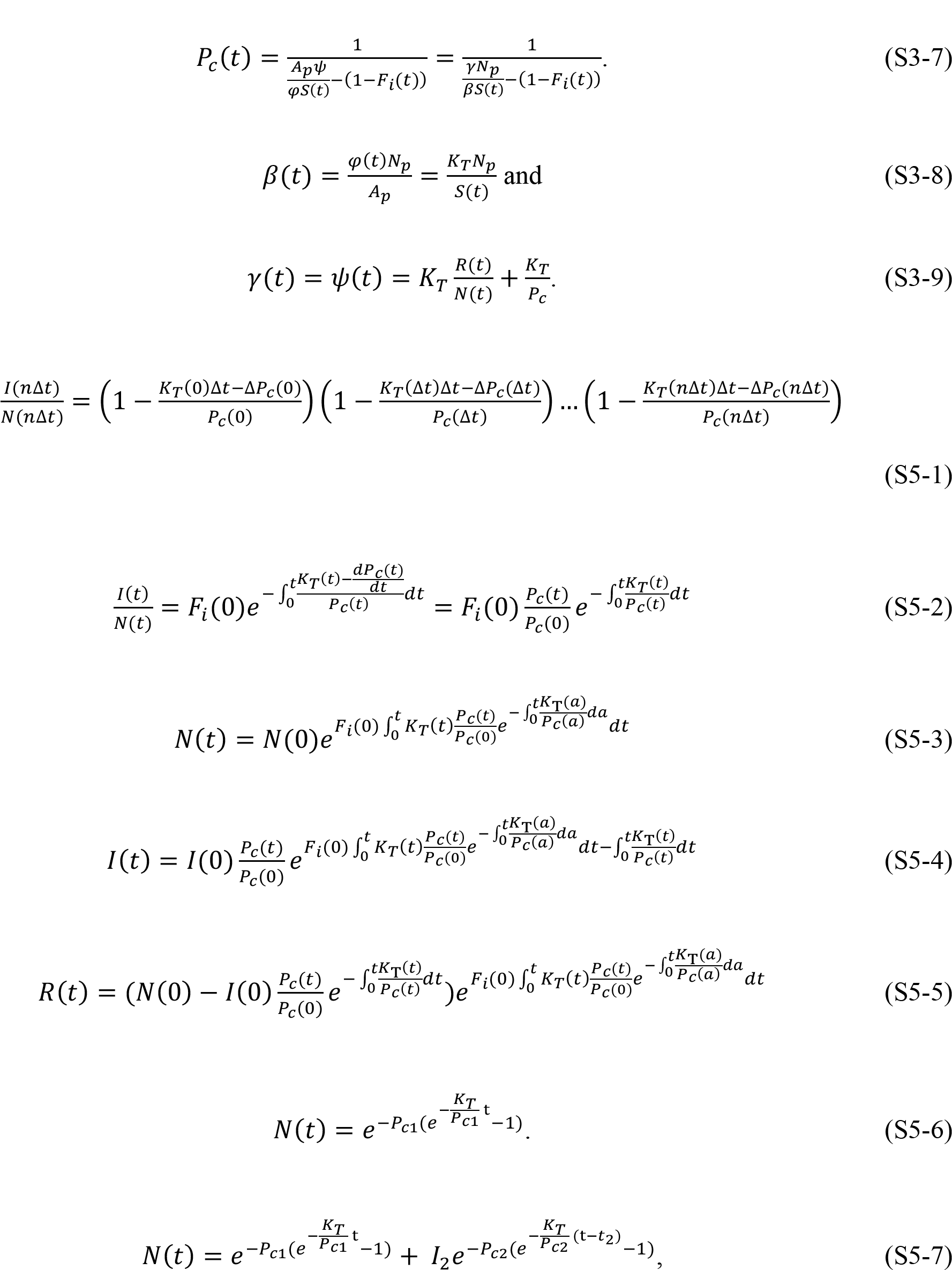
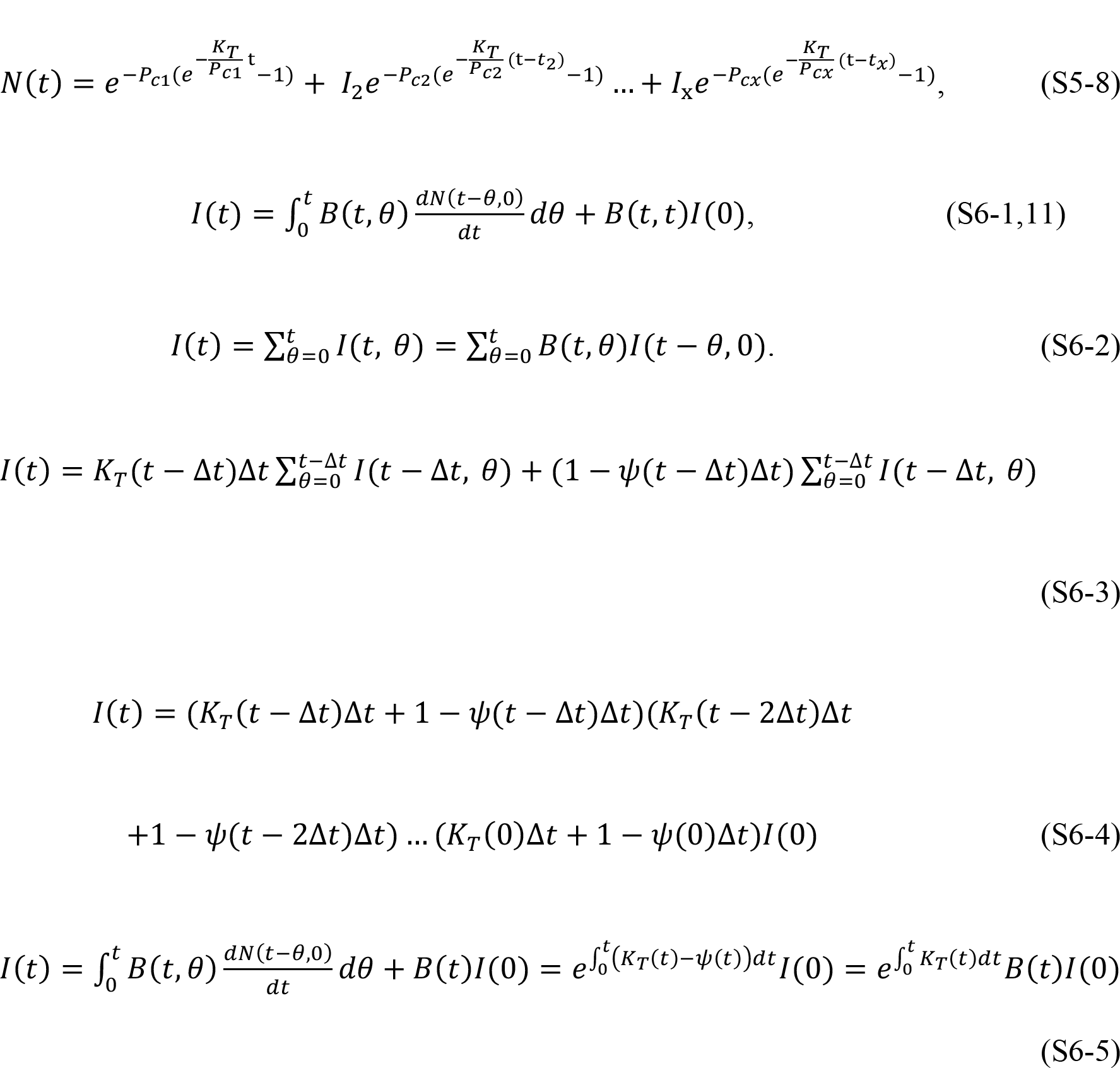

## Author contributions

T. Duclos and T. Reichert: Conceptualization, Writing – Original draft preparation, Writing – Review & Editing. T. Duclos: Methodology, Validation. T. Reichert: Data Investigation.

## Declarations of interes

none.

## Abbreviations used in this text

KMES: Kermack McKendrick integro-differential Equation Solution
RCO: Rate of Change Operator
RMM: Residential Mobility Measure
SIR: Susceptible–Infectious–Recovered

